# Sensitivity of excess mortality due to the COVID-19 pandemic to the choice of the mortality index, method, reference period, and the time unit of the death series

**DOI:** 10.1101/2021.07.20.21260869

**Authors:** Marília R. Nepomuceno, Ilya Klimkin, Dmitry A. Jdanov, Ainhoa Alustiza Galarza, Vladimir Shkolnikov

## Abstract

Estimating excess mortality is challenging. The metric depends on the expected mortality level, which can differ based on given choices, such as the method and the time series length used to estimate the baseline. However, these choices are often arbitrary, and are not subject to any sensitivity analysis. We bring to light the importance of carefully choosing the inputs and methods used to estimate excess mortality. Drawing on data from 26 countries, we investigate how sensitive excess mortality is to the choice of the mortality index, the number of years included in the reference period, the method, and the time unit of the death series. We employ two mortality indices, three reference periods, two data time units, and four methods for estimating the baseline. We show that excess mortality estimates can vary substantially when these factors are changed, and that the largest variations stem from the choice of the mortality index and the method. We also find that the magnitude of the variation in excess mortality can change markedly within countries, resulting in different cross-country rankings. We conclude that the inputs and method used to estimate excess mortality should be chosen carefully based on the specific research question.

## Introduction

Excess mortality has been considered one of the most reliable approaches for measuring the impact of the COVID-19 pandemic. The metric can be estimated by measuring the difference between mortality from all causes that is *observed* during the pandemic and mortality from all causes that would be *expected* if the pandemic had not occurred (baseline mortality). In considering deaths from all causes, excess mortality is independent of the COVID-19 testing capacity, the definition of COVID-19 deaths, and the misclassification of COVID-19 deaths on death certificates (Gill & DeJoseph, 2020; Leon et al., 2020). Moreover, the metric includes deaths that are both directly and indirectly attributable to SARS-CoV-2 (Ackley et al., 2021). As a result, estimating excess mortality has been considered the best approach for assessing and comparing the overall mortality burden due to the COVID-19 pandemic across time and space (Beaney et al., 2020).

Despite its advantages, estimating excess mortality is not a trivial task. Excess mortality estimates depend on the baseline, which can vary depending on the mortality index (death counts or rates), the number of years included in the reference period, and the method used to estimate the baseline (Németh et al., 2021; Schöley, 2021). In addition, excess mortality estimates may change depending on the time unit of the death series, which could be weekly, monthly, or quarterly. These are some of the sources of variation that could explain the differences between the estimates of excess mortality during the COVID-19 pandemic that have been provided by different authors (Bilinski & Emanuel, 2020; Islam et al., 2021; Jdanov et al., 2021; Karlinsky & Kobak, 2021; Kontis et al., 2020; Rizzi & Vaupel, 2021; The Economist DataTeam, 2020; Vestergaard et al., 2020; Wu & McCann, 2020).

Inconsistencies in excess mortality estimates can lead to poor policy decisions and affect the country rankings for excess mortality. Country comparisons are essential for assessing the efficiency of country-specific policy interventions designed to reduce the impact of the COVID-19 pandemic. However, changes in country rankings can be partially attributable to the sensitivity of each country to the inputs and methods that are used to estimate excess mortality. To date, little is known about variation in excess mortality estimates due to differences in the choice of the mortality index, the reference period, the time unit of the death data, and the method for estimating expected mortality. In addition, the question of how different combinations of these sources of variation can result in different excess mortality estimates remains open. More importantly, the magnitude of these differences has yet to be explored.

When seeking to provide excess mortality estimates, researchers should be aware that this metric depends on the expected mortality level, which can differ depending on the choices made. Thus, this study focuses on four sources of excess mortality variation: (*1*) the mortality index, (*2*) the method used to estimate the baseline, (*3*) the number of years included in the reference period, and (*4*) the time unit of the death data (weekly and monthly). We investigate to what extent excess mortality estimates depend on these sources of variation. Then, we analyze how important these factors are for each specific country and its excess mortality ranking. To do so, we calculate annual excess mortality estimates for 26 countries in 2020, and create 16 different scenarios that combine two mortality indices, four methods, three reference periods, and weekly and monthly death series to estimate the expected mortality level for 2020. Our findings indicate which of these factors, or which combinations of these factors, have a greater impact on excess mortality estimates within countries, and on the relative positions of countries. In addition, we highlight the importance of carefully choosing inputs and methods for estimating expected mortality before analyzing and drawing substantive conclusions concerning excess mortality.

## Sources of Variation in Excess Mortality Estimates

### Mortality Index

There are different ways to summarize mortality in a population, such as death rates, death counts, and life expectancy. For estimating excess mortality, death rates and death counts are the indices that are most commonly used (Bilinski & Emanuel, 2020; Faust et al., 2020; Schöley, 2021). Among the death rates researchers have used are crude death rates (CDRs) (Aburto et al., 2021; Basellini et al., 2021; Németh et al., 2021; Stokes et al., 2021), age-specific death rates (ASDRs) (Németh et al., 2021), and age-standardized death dates (SDRs) (Islam et al., 2021; Krieger et al., 2020). These indices reflect different levels and trends in mortality that may result in variation in the expected mortality level used to estimate excess mortality.

Differences in population age structures have a major influence on comparisons of CDRs and death counts between countries. CDRs express real-life mortality and population losses. However, when comparing levels of mortality across populations, it is desirable to reduce the influence of the age composition by calculating the ASDRs or the SDRs. In this study, we use CDRs and SDRs to estimate excess mortality. Being aware of differences between CDRs and SDRs is key to understanding the differences between excess mortality estimates, which can vary depending on whether an index that is affected by population age structures (CDRs) or an index that controls for differences between age compositions (SDRs) is chosen.

### Method Used to Estimate the Baseline

Using the appropriate method to estimate the baseline is crucial for achieving robust excess mortality estimates. The methods that have been used most frequently for estimating excess mortality during the COVID-19 pandemic are simple averages and regression models (Basellini et al., 2021; Bilinski & Emanuel, 2020; Modig et al., 2021; Schöley, 2021; The Economist DataTeam, 2020; Wu & McCann, 2020). In this study, we use four different methods to estimate the baseline mortality (or the expected mortality level in the absence of the pandemic) in order to evaluate the sensitivity of excess mortality estimates to the method chosen, as detailed below.

#### 1. Specific-Average

The Specific-Average is notably the most commonly used method for estimating excess mortality (Krieger et al., 2020; Modig et al., 2021; Stang et al., 2020; The Economist DataTeam, 2020; Wu & McCann, 2020). It is easy to compute and simple to understand. This method can be computed as,

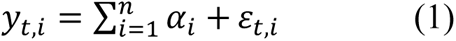

where *y_t,i_* is the expected death rate for the year *t* and the fraction of the year *i* that can be a week or a month, *n* is the total number of weeks or months in the year *t*, *α_i_* are the coefficients specific for the week or month *i*, and *ε_t,i_* are the residuals. We model the expected death rates using ordinary least squares linear regression.

#### 2. Specific-Average with Trend

The Specific-Average method does not consider mortality trends. Some researchers have already noted the importance of including time trends in their models for estimating excess mortality (Basellini et al., 2021; Karlinsky & Kobak, 2021; Németh et al., 2021; Schöley, 2021). Thus, the second model we employ includes linear time trends. To take these trends into account in building the baseline, we added to Equation (1) a term that accounts for the annual trend in death rates, as

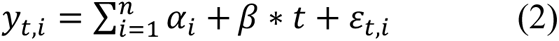

where *β* is the coefficient associated with the linear trend.

#### 3. Harmonic with Trend

The third method is a variant of the Serfling model (Serfling, 1963) which considers the seasonality of deaths over a year. This method and its variants are commonly used to estimate excess mortality due to influenza (e.g. Simonsen et al., 2005; Thompson et al., 2009). In this study, we employ the following model,

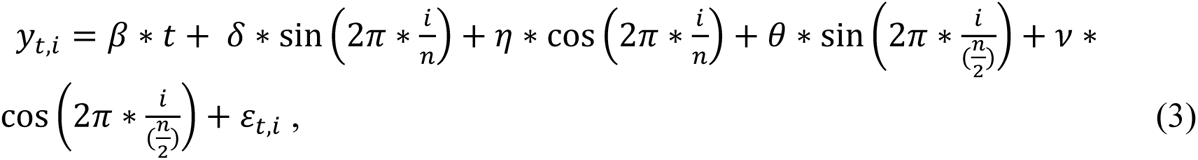

where *y_t,i_* is the expected death rate for the year *t* and the fraction of the year *i*, *i* is the *i*^th^ week or month of the year *t*, *n* is the total number of weeks or months over the year *t*, *β* is the coefficient associated with the trend, *δ*, *η*, *θ*, and *v* are the coefficients associated with the seasonality, and *ε_t,i_* are the residuals. *y_t,i_* are the expected death rates of a linear model estimated by ordinary least squares.

#### 4. Specific-Trend

The fourth method we employ in this study is used less frequently to estimate excess mortality (Németh et al., 2021). It considers the linear time trend of the fraction of the year *i* that can be a week or a month, and can be computed as,

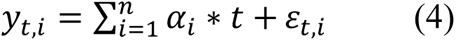

where *y_t,i_* is the expected death rate for the year *t* and the fraction of the year *i*, *n* is the total number of weeks or months in the year *t*, *α_i_* are the coefficients specific for the fraction of the year *i*, and *ε_t,i_* are the residuals.

### Reference Period

The reference period, defined as the number of previous years included in the baseline, is also a source of variation across excess mortality estimates. The goal of the baseline is to provide a reference level of mortality that reflects as accurately as possible recent mortality patterns that would have occurred in the absence of the pandemic. Many of the studies on excess mortality due to the COVID-19 pandemic have arbitrarily chosen the reference period of 2015-2019 (Bilinski & Emanuel, 2020; Krieger et al., 2020; Schöley, 2021). However, we believe that the choice of the number of years included in the baseline should be based on certain considerations. For instance, for countries that have experienced steeper declines in mortality in the early years of the last decade than in the final years of the decade, the expected level of mortality for 2020 will be lower than that based on more recent years if 2010-2019 is defined as the reference level instead of a more recent period, such as 2017-2019. Thus, the choice of the reference period can affect excess mortality estimates.

In addition, depending on the reference period considered in the baseline, some previous epidemic years may be included in the expected mortality level. In 2015, for instance, European countries experienced a severe influenza epidemic, which resulted in higher levels of mortality in 2015 than in the previous winter season (Fedeli et al., 2017; Nielsen et al., 2019). Thus, if 2015 is included in the reference period, the elevated mortality in that year will increase the mortality level of the baseline. This example highlights the importance of carefully choosing the years included in the reference period used to estimate excess mortality.

To evaluate the sensitivity of excess mortality estimates to changes the reference period, we consider three different periods: (*i*) 2010-2019, (*ii*) 2015-2019, and (*iii*) 2017-2019. We have chosen these three reference periods because they reflect, respectively, the mortality level based on a long time series (2010-2019), the mortality level based on a more recent mortality pattern (2015-2019), and the mortality level in the absence of the acute epidemic of 2015.

### Data Time Unit (weekly and monthly death series)

Weeks and months are the two time units of the death data that are used most frequently to estimate excess mortality (Jdanov et al., 2021; Karlinsky & Kobak, 2021). The use of weekly data provides more precise mortality estimates and the opportunity to obtain information about recent mortality shocks with a minimal delay. However, for several countries, weekly data are not easily available, and monthly data have been used instead. Among the potential pitfalls of using monthly data is that in addition to being a slower way to monitor changes in mortality during a mortality crisis, it provides a smoother time series than using weekly data would. However, little is known about the comparability of excess mortality estimates derived from weekly and monthly data. In this study, we employ weekly and monthly death series to estimate excess mortality, and, in turn, to investigate how the choice of the data time unit affects excess mortality.

## Sensitivity analysis

Having defined the sources of variation in excess mortality estimates, we evaluate to what extent excess mortality estimates depend on the combination of (*1*) the mortality index used and (*2*) the method employed to estimate the baseline. We hypothesized that different combinations of a specific mortality index with a given method could result in different excess mortality estimates. Therefore, we have created eight scenarios in which we apply the four methods to each mortality index. To investigate the sensitivity of the mortality index and the method, we assume, in these eight scenarios, that the reference period is 2015-2019, and the data time unit is weekly, as shown in Table 1.

**Table 1.** Scenarios for the sensitivity analysis by varying the mortality index and the method.

| Scenario | Mortality Index | Method | Reference Period | Time Unit |
| --- | --- | --- | --- | --- |
| Scenario 1 | SDR | Specific-Average | 2015-2019 | Weeks |
| Scenario 2 | SDR | Specific-Average with Trend | 2015-2019 | Weeks |
| Scenario 3 | SDR | Harmonic with Trend | 2015-2019 | Weeks |
| Scenario 4 | SDR | Specific-Trend | 2015-2019 | Weeks |
| Scenario 5 | CDR | Specific-Average | 2015-2019 | Weeks |
| Scenario 6 | CDR | Specific-Average with Trend | 2015-2019 | Weeks |
| Scenario 7 | CDR | Harmonic with Trend | 2015-2019 | Weeks |
| Scenario 8 | CDR | Specific-Trend | 2015-2019 | Weeks |

Source: Author's elaboration.

Then, to investigate the impact of the reference period on excess mortality estimates, we have created four additional scenarios, as shown in Table 2. In Scenarios 9-12, we vary the reference period by using 2010-2019 and 2017-2019 instead of by using 2015-2019, as in Table 1. We keep constant the method and the time unit of the death series. Scenarios 9-12 are built for both SDRs and CDRs.

**Table 2.** Scenarios for the sensitivity analysis by varying reference period.

| Scenario | Mortality Index | Method | Reference Period | Time Unit |
| --- | --- | --- | --- | --- |
| Scenario 9 | SDR | Specific-Average with Trend | 2010-2019 | Weeks |
| Scenario 10 | SDR | Specific-Average with Trend | 2017-2019 | Weeks |
| Scenario 11 | CDR | Specific-Average with Trend | 2010-2019 | Weeks |
| Scenario 12 | CDR | Specific-Average with Trend | 2017-2019 | Weeks |

Source: Author's elaboration.

To evaluate the magnitude of the variation in excess mortality by varying the reference period from 2015-2019 to 2010-2019 and 2017-2019, the scenarios presented in Table 2 should be compared with Scenarios 2 and 6 from Table 1. Scenario 2 for SDRs and Scenario 6 for CDRs employ the same method and time unit of Scenarios 9-12, but for the 2015-2019 reference period.

Table 3 shows four additional scenarios designed to evaluate the impact on excess mortality of changes in the data time unit. In Scenarios 13-16, we vary the time unit of the death series from weekly to monthly. Unlike in the previous scenarios, we consider weeks 1-52 in leap week years in order to compare monthly with weekly death series, and to avoid potential disagreement between these two time units derived from the 53^rd^ week in leap week years,

**Table 3.** Scenarios for the sensitivity analysis by the data time unit.

| Scenario | Mortality Index | Method | Reference Period | Time Unit |
| --- | --- | --- | --- | --- |
| Scenario 13 | SDR | Harmonic with Trend | 2015-2019 | Weeks (1-52) |
| Scenario 14 | SDR | Harmonic with Trend | 2015-2019 | Months |
| Scenario 15 | CDR | Harmonic with Trend | 2015-2019 | Weeks (1-52) |
| Scenario 16 | CDR | Harmonic with Trend | 2015-2019 | Months |

Source: Author's elaboration.

In Table 3, we combine monthly and weekly (weeks 1-52) data with the Harmonic with Trend method. We have chosen to employ this method because the use of monthly or weekly data combined with the Specific-Average or Specific-Average with Trend methods should, theoretically, provide equal excess mortality estimates, given that these methods are mathematically equivalent with respect to the time unit (Karlinsky & Kobak, 2021). However, since this is not the case for the Harmonic with Trend method, we believe that differences in excess mortality may emerge when this method is combined with different time units.

To investigate the magnitude of variations in excess mortality estimates by using monthly instead of weekly data, we compare the scenarios in Table 3 within each mortality index. For instance, the magnitude of the variation in excess mortality by using monthly instead of weekly data for the SDRs, the Harmonic with Trend method, and the 2015-2019 reference period, is equal to the difference between the excess mortality levels estimated by Scenario 13 and Scenario 14.

## Data

We used data from the Short-Term Mortality Fluctuations (STMF) data series (Jdanov et al., 2021), which is a new component of the Human Mortality Database (HMD) (Barbieri et al., 2015). We draw data from 26 countries/regions. Of these 26 countries/regions, 23 have complete series lasting from 2010 to 2020, including Austria, Belgium, Denmark, England and Wales, Estonia, Finland, France, Hungary, Israel, Latvia, Lithuania, the Netherlands, Norway, Poland, Portugal, Republic of Korea, Scotland, Slovenia, Slovakia, Spain, Sweden, Switzerland, and Taiwan. In Italy, New Zealand, and the United States, the time series start from 2011, 2011 and 2015, respectively.

Weekly deaths are directly drawn from the STMF data series, which follow the ISO 8601-2004 guidelines. Monthly deaths are collected from the national statistical offices. The figures are published by these offices, or they are requested by the STMF team. Monthly deaths are available for the 26 countries. Appendix A presents the list of monthly data sources by country.

Population exposures by week are retrieved from the STMF data series. Following the STMF approach, the monthly population exposures were calculated as the annual exposures divided by 12 (Jdanov et al., 2021).

From the weekly and monthly data, we calculate annual mortality rates for all ages and men and women together. The HMD core provides annual death rates from 2010 to 2018 for all the countries considered in this study, except Hungary (2010-2017), Israel (2010-2016), New Zealand (2010-2013), Slovenia (2010-2017), and Slovakia (2010-2017). The available HMD annual death rates were forecasted up to 2020 (Jdanov et al., 2021). However, since the STMF data series are preliminary, some deaths may not be included in the weekly series. In addition, because the STMF data series use ISO weeks (each week has seven days), the death rates from the STMF might be slightly different from the annual death rates derived from the HMD core. Therefore, we compare annual death rates from both the STMF data series and the HMD core data, and thus adjust for any differences that might emerge, while assuming that the HMD core data represent the gold standard.

To make the annual death rates derived from weekly data comparable across leap week years (years with 53 weeks) and non-leap week years (years with 52 weeks), we calculate baseline mortality levels based on weeks 1-52 and assume that the baseline for week 52 is equal to that of week 53. The monthly death counts were adjusted as well. Months in leap years have, on average, 30.50 days (366/12), while the average number of days per month in non-leap years is 30.42 (365/12). We assume that the average number of days in both leap and non-leap years is 30.44 days (365.25/12). Thus, we multiply the number of deaths in each month by the ratio between 30.44 and the actual number of days in each month. Then, to make sure that the total number of deaths in a year did not change, we adjust for any eventual differences in the annual total number of deaths before and after considering that the average number of days in a month is 30.44.

To age-standardize the crude death rates, we use the European Population Standard of 2013 (European Comission, 2013), and apply the methodology developed by Klimkin et al. (2021).

## Results

Figure 1 shows the levels and trends in observed death rates between 2010-2019, and highlights variations in the estimated levels of expected mortality in 2020 for France, the US, Belgium, Hungary, and Poland. The figure shows temporal changes in the observed CDR and SDR values up to 2019, and two values of the expected rates depending on the method used to estimate the baseline (Specific-Average and Specific-Average with Trend) in 2020.

**Figure 1:**
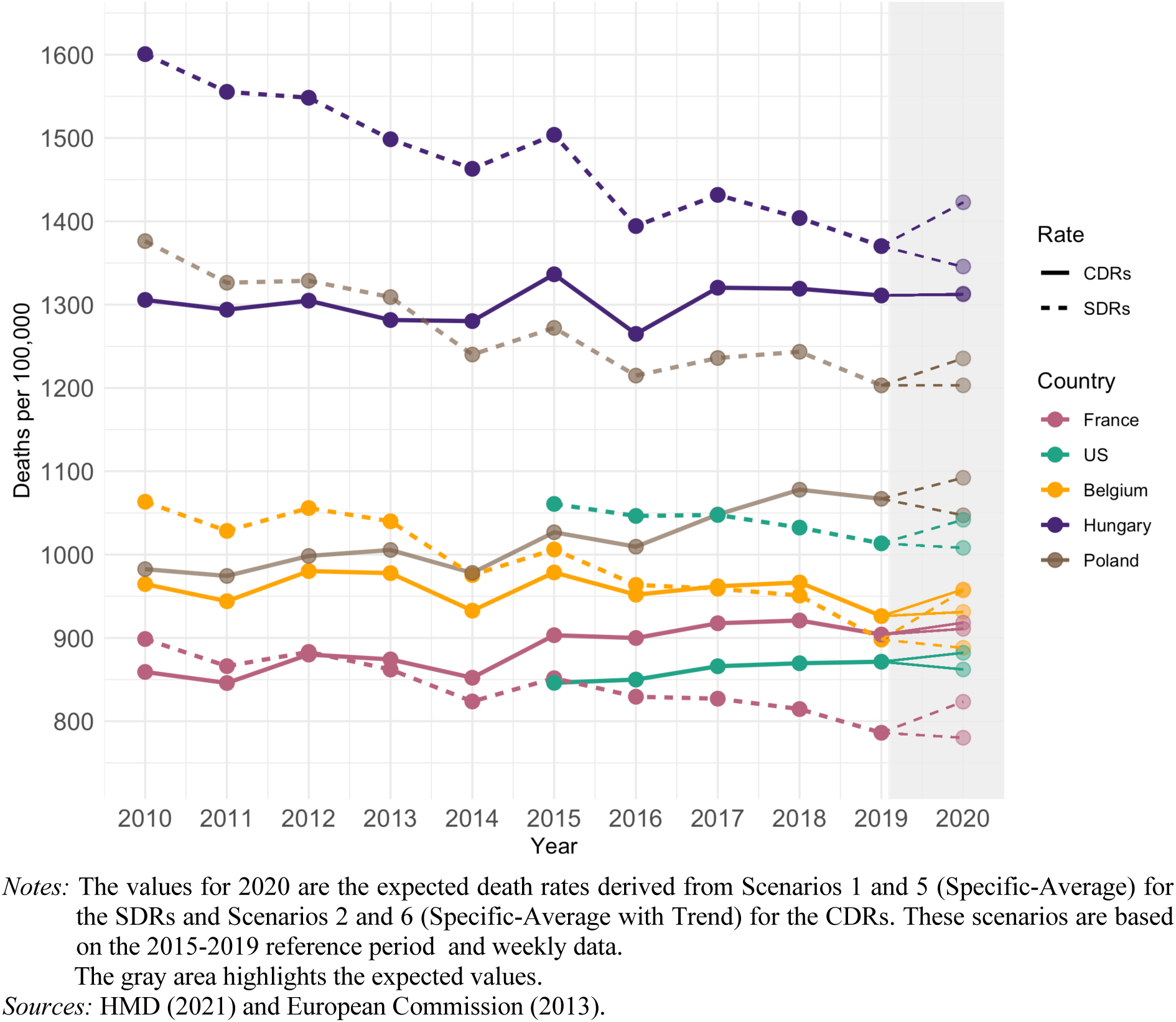
Observed crude death rates (CDRs) and age-standardized death rates (SDRs) in France, the US., Belgium, Hungary, and Poland in 2010-2019, and their expected values in 2020.

Figure 1 introduces our later analysis by showing that both the levels and the trends change considerably with the mortality index used between 2010 and 2019. In the US, Poland, and Hungary, the SDR is found to be much higher than the CDR. In 2015, for instance, the SDR was 25% higher than the CDR in the US, and was 13% higher than the CDR in Hungary. Figure 1 also reveals a striking level of variation in the trends of the two indices. In France and in the US, the CDR increases over time, while the SDR decreases. In Hungary, the SDR decreases, while the CDR remains nearly stable between 2017 and 2019. The differences in the levels and trends in the CDRs and the SDRs depending on the methods used resulted in variation in the expected level of mortality in 2020, as the gray area of Figure 1 shows.

### Sensitivity of excess mortality estimates to mortality index and method

Figure 2 illustrates in greater detail the impact on excess mortality of the baseline variations presented in Figure 1. Figure 2 displays the excess mortality rates by varying the method used to calculate the baseline for both the CDR and the SDR. The figure demonstrates that different combinations of the mortality index and the method used result in different excess mortality estimates for all countries, as was already shown in Figure 1. Figure 2 illustrates the impact of the method used on the excess mortality estimates for each mortality index. For both the CDR and the SDR, the three methods that consider time trends (Specific-Average with Trend, Harmonics with Trend, and Specific-Trend) estimate similar excess mortality rates for all countries, while the Specific-Average is the method that provides excess mortality estimates that tend to result in greater variation. For the CDR, the Specific-Average method can produce higher or lower levels of excess mortality than the other methods. In Italy, Poland, and the US, the Specific-Average method provides higher excess mortality rates than the other methods; while in Sweden, Israel, and Norway, the Specific-Average method provides lower excess mortality rates than the other methods. For the SDR, the Specific-Average method systematically provides lower excess mortality rates for all countries than the methods that account for linear trends. In Lithuania, for instance, the excess mortality for SDR estimated by the Specific-Average method is nearly four times lower than that derived from the other methods. Appendix B, Table 1B shows the excess mortality values presented in Figure 2.

**Figure 2.**
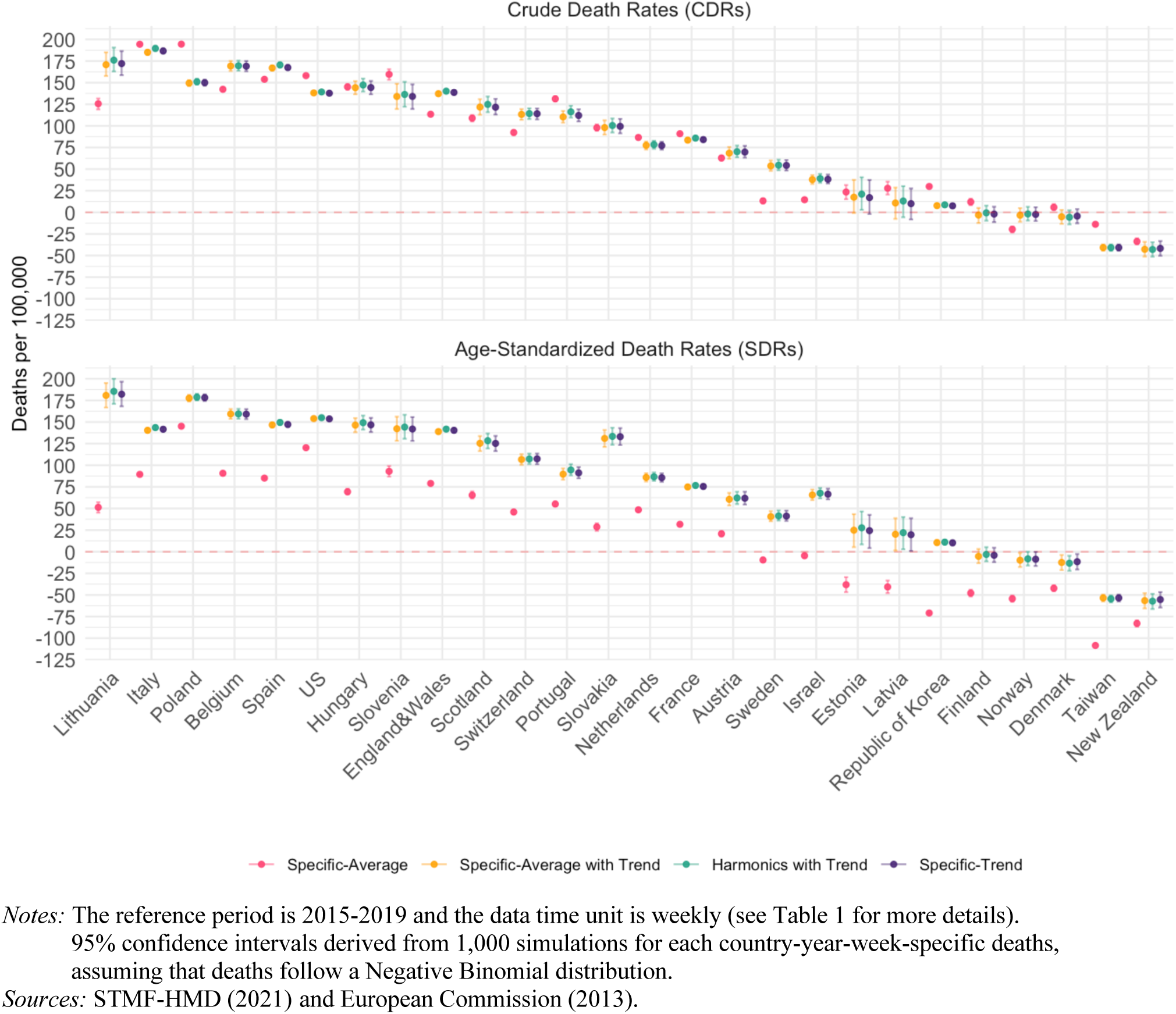
Excess mortality estimates by varying the method for each mortality index and by country, 2020.

To isolate the effect of the mortality index on excess mortality estimates and to highlight the magnitude of its variation, Figure 3 presents the absolute difference between the excess mortality rates estimated by using the CDR and the SDR across the four methods. Figure 3 shows substantial differences between excess mortality estimates when the mortality index is varied within each method for all countries. The greatest differences emerge when the Specific-Average method is employed for all countries, except for Israel. In Italy and the Republic of Korea, variations in excess mortality rates (per 100,000) due to the use of the CDR or the SDR combined with the Specific-Average method are above 100 deaths. In Israel, the Specific-Average is the method that results in the lowest variation in excess mortality when the mortality index varies. For all countries, the differences in the excess mortality estimates depending on the mortality index used are smaller for the methods that account for trends. For example, in Hungary, there is virtually no variation in the excess mortality rates depending on the use of the CDR or the SDR for all baseline mortality levels that account for the trend.

**Figure 3.**
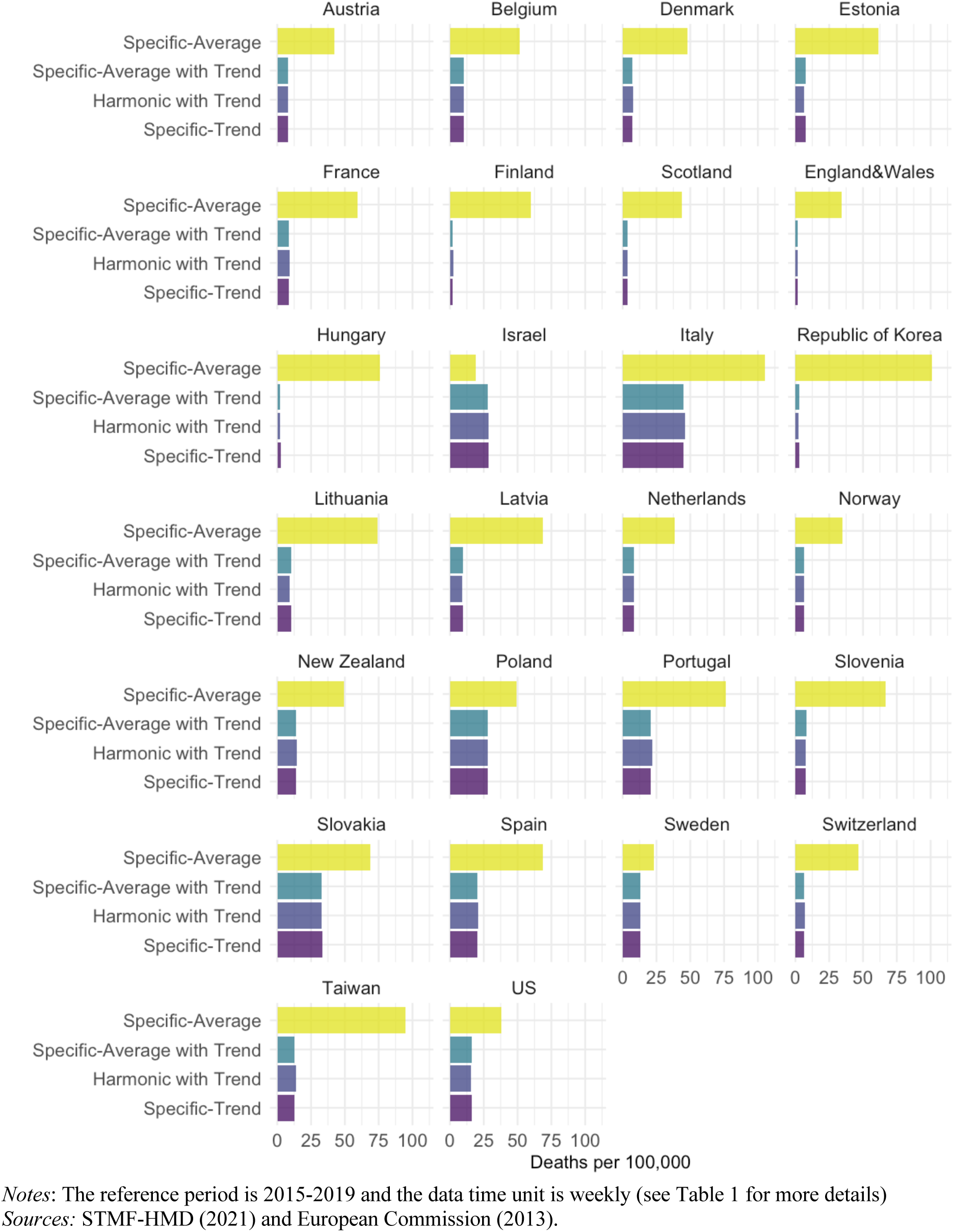
Absolute differences between the excess CDR and the excess SDR values for each method used to estimate excess mortality.

### Sensitivity of excess mortality estimates to the reference period and the data time unit

Figure 4 shows that excess mortality rates can vary by changing the reference period for each mortality index using the Specific-Average with Trend method (Scenarios 9-12). This figure suggests that when the reference period is longer, the excess mortality rates are lower for most countries. However, the excess mortality estimates depend on the trend of the mortality index in the chosen reference period. In Belgium, for instance, where there was a steeper decline in the SDR between 2015 and 2019 than in the 2010-2019 reference period (Figure 1), the excess mortality rate derived from the 2010-2019 reference period was 20% lower than that derived from the 2017-2019 reference period. Poland experienced a different SDR trend than Belgium (Figure 1). In Poland, the decline in the SDR was steeper between 2010 and 2019 than it was between 2015 and 2019 (Figure 1). As a result of this trend, excess mortality rates appear to be higher in Poland when the 2010-2019 period is considered rather than the 2015-2019 period. Appendix B, Table 2B presents all of the excess mortality values displayed in Figure 4.

**Figure 4.**
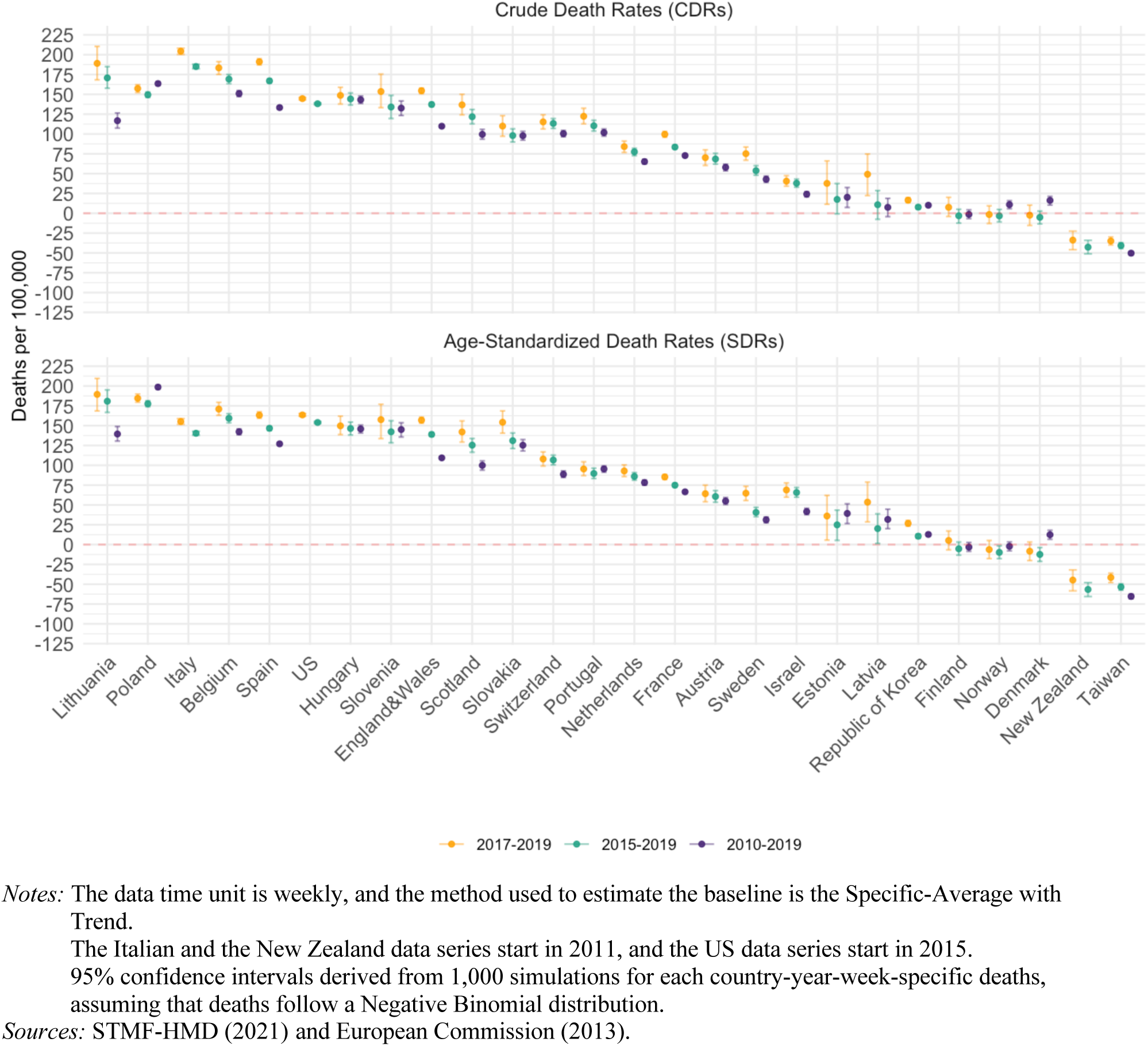
Excess mortality estimates by varying the reference period for each mortality index and country, 2020.

Figure 5 shows the magnitude of the differences in the estimated excess mortality when the reference period for each death rate is varied. The magnitude of the differences in excess mortality rates changes substantially within countries when the reference period is changed, ranging from 0.1 to 55 deaths. Moreover, the magnitude of the differences within countries varies when the mortality index is combined with the reference period. By changing the reference period from 2010-2019 to 2015-2019, the differences between the excess mortality rates (per 100,000) in Lithuania are 54 for the CDR and 40 for the SDR. For Portugal, Figure 5 presents a different pattern: for the SDR, the differences are very small depending on whether the 2010-2019 or the 2017-2019 reference period is used instead of the 2015-2019 reference period; while for the CDR, the variation in excess mortality rates is higher when the 2017-2019 reference period is used instead of the 2015-2019 reference period.

**Figure 5.**
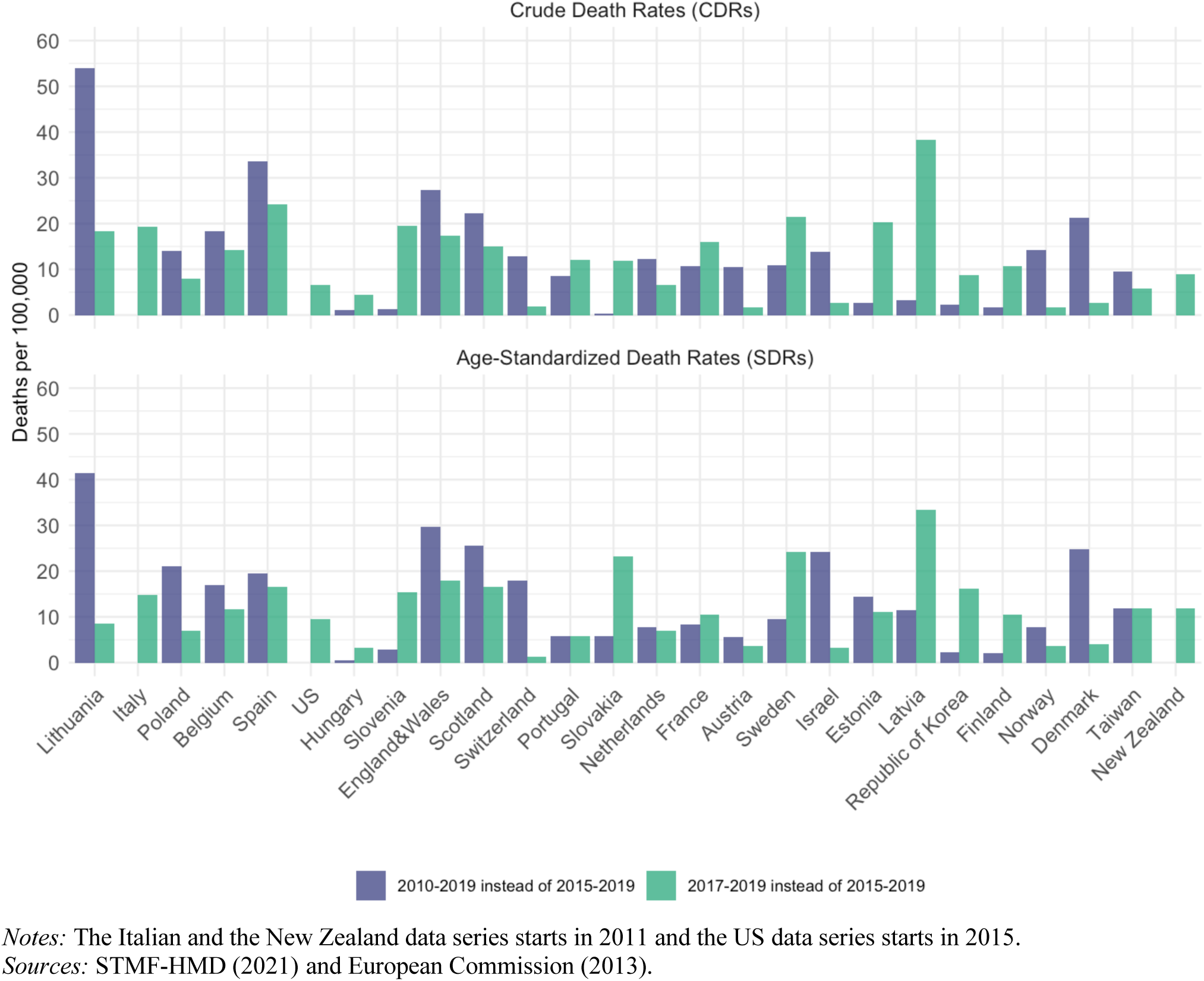
Absolute difference in excess mortality estimates by varying the reference period for each mortality index and country, 2020.

Figure 6 compares excess mortality depending on whether weekly or monthly death series are used in combination with the Harmonic with Trend method (Scenarios 13-16). The excess mortality estimates are very similar for both data time units across all countries and for both rates.

**Figure 6.**
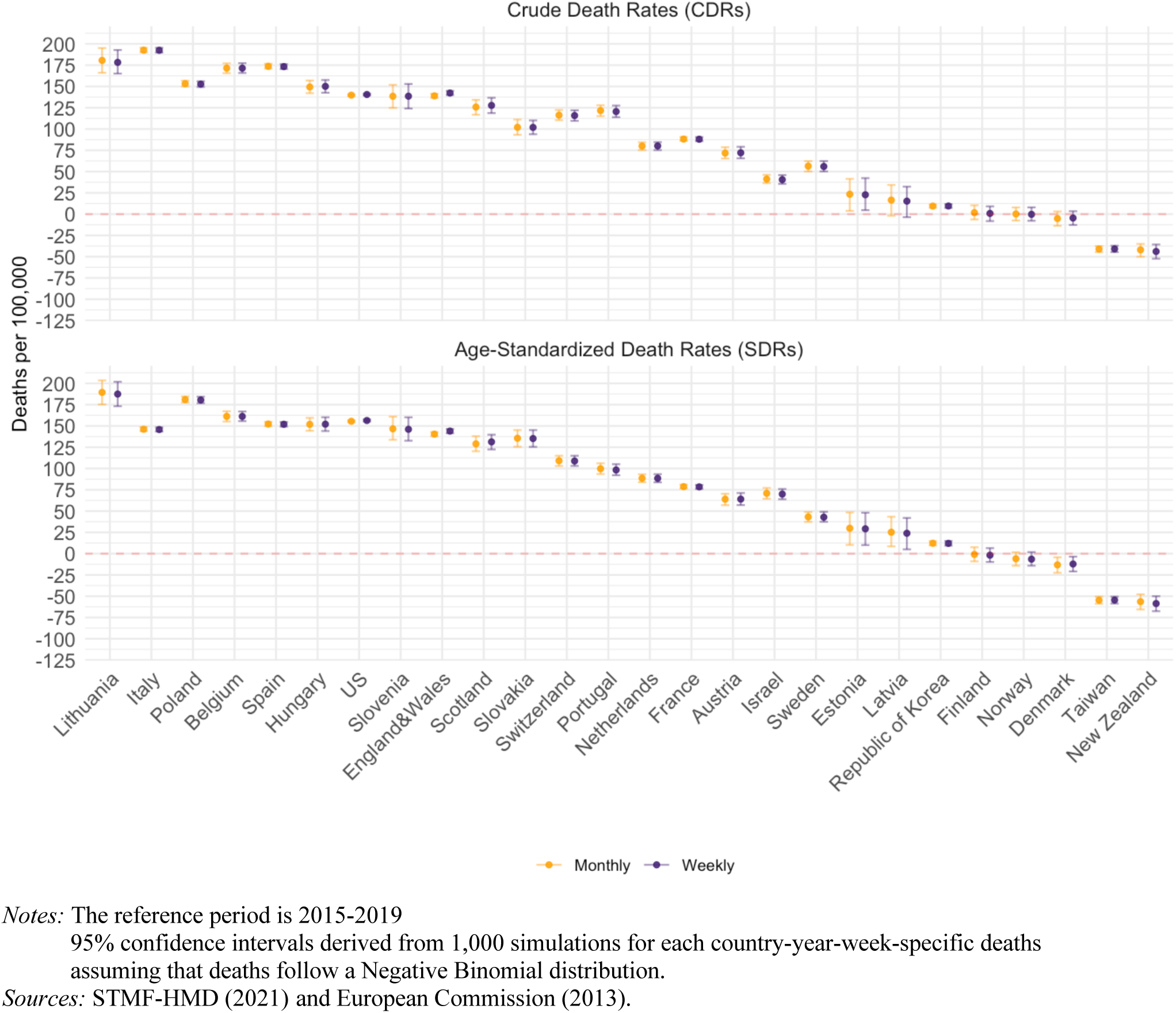
Excess mortality estimates by varying the time unit of the death series for each mortality index and country, 2020.

Figure 7 complements Figure 6 by showing the magnitude of the excess mortality differences by using monthly instead of weekly data. The differences are very small across all countries and for both rates (below 3.5 deaths per 100,000 person-years). In France, for instance, the change in excess mortality rates is below 0.5 deaths. The largest difference is in England and Wales, where the variation in excess mortality rate (per 100,000) is about 3.5 deaths.

**Figure 7.**
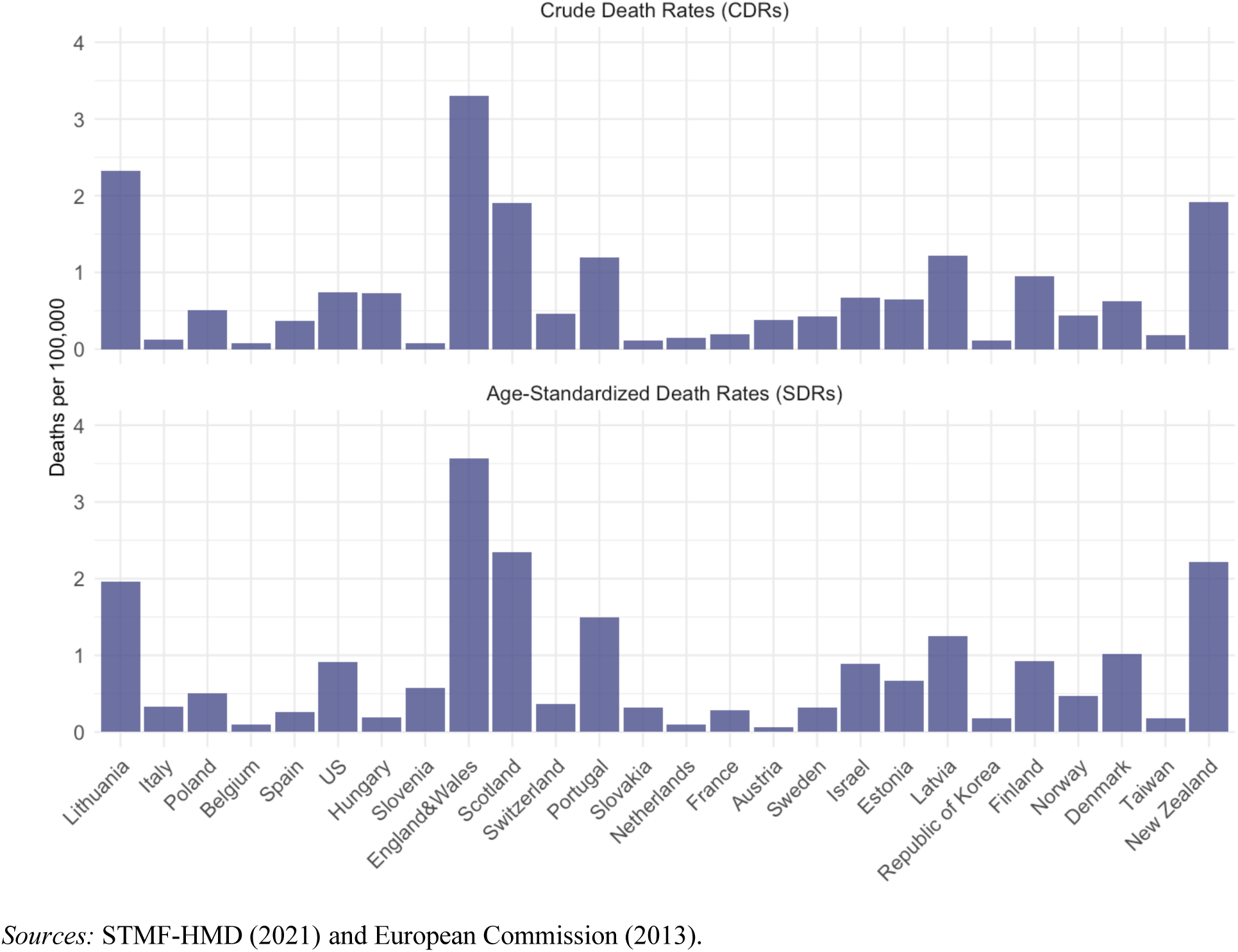
Absolute difference in excess mortality estimates by using monthly instead of weekly data, for each mortality index and country, 2020.

As we mentioned in the Sensitivity Analysis section, theoretically, excess mortality estimated with the Specific-Average with Trend method should not vary when the data time unit is changed. To empirically test this theoretical claim, we compare the results shown in Figure 7 with excess mortality rates when the Specific-Average with Trend method is employed instead of the Harmonic with Trend method. Figure 1C in Appendix C presents this comparison. As expected, Figure 1C shows that variations in excess mortality are generally lower when the Specific-Average Trend method is used. However, in this comparison, we use a reference period that includes the 2015 leap year, which can lead to discrepancies between monthly and weekly data. Thus, Figure 1C also presents the variation in excess mortality due to the data time unit when the Specific-Average Trend method is combined with the 2017-2019 reference period. This latter combination results in virtually no differences between excess mortality estimates depending on whether monthly or weekly data are used. Thus, Figure 1C suggests that both the method employed and the use of leap years in the reference period lead to in variations in excess mortality derived from monthly or weekly death series.

### Country Ranking

Figure 8 presents Spearman’s correlation coefficients between excess mortality rankings for Scenarios 1-12 (S1-S12) across the 26 countries. Scenarios 13-16, which evaluate variations in excess mortality due to the choice of the data time unit, were not included in this analysis because they do not consider the 53^rd^ week of leap week years in the baseline, while other scenarios account for that week. This figure highlights the similarities and the differences in the countries’ excess mortality rankings estimated for each scenario presented in Tables 1-2. Darker blue hues indicate a stronger correlation between the excess mortality pair comparisons.

**Figure 8.**
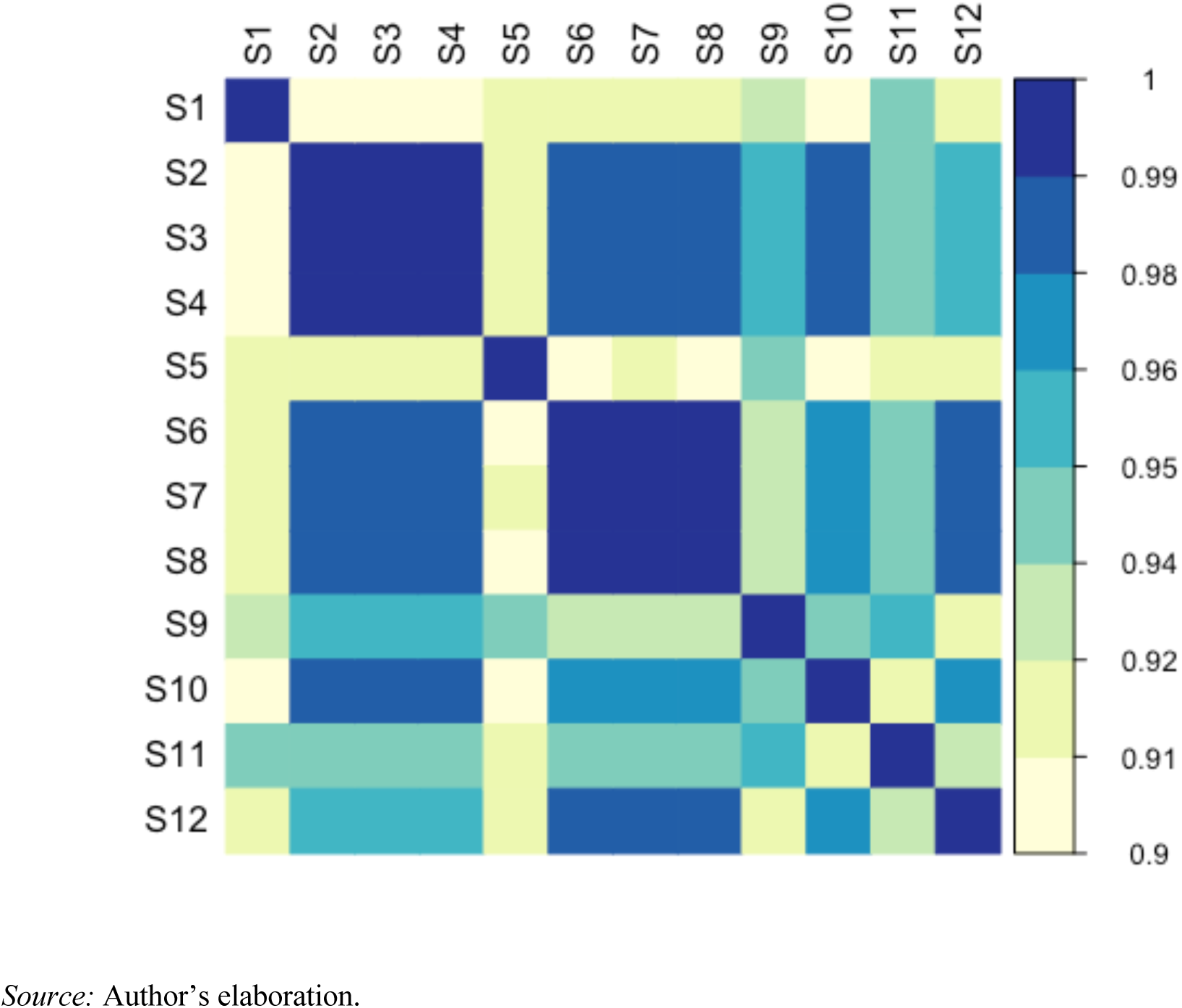
Spearman’s correlation coefficients between excess mortality rankings for the 12 scenarios across the 26 countries, 2020.

The Spearman’s correlation coefficients for different pairs of excess mortality are above 0.9, which indicates a high degree of correlation between the excess mortality estimates (Figure 8). A lower degree of correlation is observed in pair comparisons that involve Scenario 1 (S1) and Scenario 5 (S5). Scenario 1 for the SDR and Scenario 5 for the CDR, employ the only method that does not account for the trend, the Specific-Average method (see Table 1 for more details). This finding highlights the impact of choosing a method with or without a linear trend on country rankings. Discrepancies in rankings disagreements are also observed when scenarios that consider the 2010-2019 reference period (S9 and S11) are combined with the other scenarios. On the other hand, the highest correlation in rankings is observed in scenarios in which the method accounts for the trend term. This finding suggests that for a given mortality index and reference period, employing different methods that consider linear trends provides very similar country rankings.

In addition to the Spearman’s correlation coefficients shown in Figure 8, Table 4B in Appendix B presents the country rankings for excess mortality from the highest to the lowest for each country and across all scenarios. This table shows some country-specific changes in the country rankings. For instance, when moving from Scenario 1 to Scenario 2, which consider he Specific-Average method and the Specific-Average with trend method, respectively, Lithuania rises from the 11^th^ to the first position, while Slovakia rises from the 15^th^ to the 10^th^. For Italy, Table 4B shows a marked change in the ranking when CDR (Scenarios 6-8) is employed instead of SDR (Scenarios 2-4): i.e., the country rises to the first position when CDR is used.

## Discussion

We investigated the sensitivity of excess mortality estimates in 2020 to the choice of the mortality index, the method, the reference period, and the time unit of the death series in 26 countries/regions. Our results showed when these factors changed, the excess mortality estimates varied substantially, and the magnitude of these variations changed markedly within countries, which resulted in changes in the country rankings.

The choice of the mortality index was found to be one of the main sources of excess mortality variation within countries. We used two mortality indices that provided excess mortality estimates with and without the influence of the population age structure. We showed that both the levels and the trends differed substantially for the CDR and the SDR, which provided diverse baseline mortality levels, and which, in turn, led to variations in the excess mortality estimates.

In the context of population aging, whether the CDR or the SDR is used is crucial for country comparisons. Differences between population age structures are confounding factors when the goal is to compare the populations’ mortality levels (Preston et al., 2001). A higher proportion of older adults combined with the steep age gradient in COVID-19 mortality (Goldstein & Lee, 2020) (considering that excess deaths were largely driven by deaths due to SARS-CoV-2) resulted in higher crude death rates for older than for younger populations. Italy is a good example of this phenomenon, because it has one of the oldest populations in the world (Gesano & Strozza, 2011; Mazzola et al., 2016; Murphy, 2017). Our findings showed that the magnitude of the variation in the excess mortality rate (per 100,000) for Italy varied between 45 and 105 deaths depending whether the CDR or the SDR was used. Thus, there were striking variations in the Italian position in country rankings, with Italy rising to the top highest position in the ranking when the CDR was used. We therefore conclude that differences in countries’ age compositions have a relevant impact on country comparisons of excess mortality.

The method used to estimate the baseline also appears to be an important source of variation in excess mortality. We showed that methods that considered linear trends provided similar excess mortality rates for both the CDR and the SDR. However, we also found that the Specific-Average method produced the lowest excess mortality rates across all countries for the SDR, which is in line with the findings of Schöley (2021). However, while a similar pattern was observed across all countries, the magnitude of the variation in excess mortality rates due to the choice of a method that did or did not account for trends changed for each country, ranging from about 130 deaths in Lithuania to 26 deaths in New Zealand. On the other hand, we found no similar patterns across countries when the CDR was combined with the Specific-Average method. This combination produced the lowest excess mortality estimates in nine out of 26 countries, including Sweden, England and Wales, and Israel; while in the other 17 countries, it produced the highest excess mortality estimates. In addition, our findings provided further evidence that the choice of the method matters to understand variations in excess mortality, as we found that the magnitude of these variations was country-specific, and depended on the selection of the mortality index.

In addition, we observed that the choice of the reference period also matters when estimating excess mortality. In contrast with previous research on excess mortality in which the reference period was arbitrarily chosen the reference period (Bilinski & Emanuel, 2020; Karlinsky & Kobak, 2021; Schöley, 2021), we showed that there were important variations in excess mortality estimates depending on the reference period chosen. Our findings indicated that for most countries, longer reference period resulted in lower excess mortality. More importantly, we highlighted the relevance of the trend of the mortality index in the chosen reference period, especially when methods for estimating the baseline accounted for the trends.

Furthermore, the analysis found that the data time unit of the death series was the factor that was associated with the smallest variations in excess mortality estimates. Our findings showed that the use of monthly or weekly data resulted in very similar excess mortality rates across all countries. However, the impact of the data unit on excess mortality estimates depended on both the method used and the presence of leap years in the baseline mortality level. The choice of a method that is equivalent regarding the time unit, such as the Specific-Average or the Specific-Average with Trend (Karlinsky & Kobak, 2021), reduced the impact of the data unit chosen on variations in excess mortality. Moreover, we found that when these methods were combined with a reference period that did not include leap years in the baseline, the variations were reduced, and there were virtually no changes in excess mortality depending on whether monthly or weekly data were used. Nonetheless, caution is needed when excess mortality levels estimated from different data time units are compared, as we showed by employing the Harmonic with Trend method combined with the 2015-2019 reference period.

To conclude, this study showed that when estimating excess mortality, all of the inputs and the methods used should be chosen carefully. Excess mortality estimates depend on the choice of the mortality index, the method, the reference period, and the data time unit. In addition, we found evidence that certain combinations of these choices can result in substantial variations in excess mortality. Thus, since estimating excess mortality is key to measuring the full impact of the COVID-19 pandemic on mortality and to guiding health policies, we emphasize that when considering the conclusions of analyses of excess mortality, all of the factors used to estimate the baseline mortality should be carefully considered.

## Data Availability

The data used in this study are available in the Human Mortality Database

https://www.mortality.org/

## Appendix A List of monthly data sources by country

**Austria**. Statistik Austria: Deceased by demographic criteria. Available at https://statcube.at/statistik.at/ext/statcube/jsf/tableView/tableView.xhtml (accessed on 06.03.2021)

**Belgium**. STATBEL: Number of deaths per day, sex, age, region, province, district 2009-2021. Available at https://statbel.fgov.be/en/open-data/number-deaths-day-sex-district-age (accessed on 03.03.2021)

**Denmark**. Statistics Denmark: Deaths by day of death and month of death (2007-2020). Available at https://www.statbank.dk/statbank5a/SelectVarVal/Define.asp?MainTable=DODDAG&PLanguage=1&PXSId=0&wsid=cftree (accessed on 03.03.2021)

**England & Wales**. Office for National Statistics: Monthly figures on deaths registered by area of usual residence, 2006-2020. Available at https://www.ons.gov.uk/peoplepopulationandcommunity/birthsdeathsandmarriages/deaths/datasets/monthlyfiguresondeathsregisteredbyareaofusualresidence (accessed on 03.03.2021)

**Estonia.** Statistics Estonia: Dataset:RV04: Preliminary data of registration of deaths by month and county of the registration. Available at https://andmed.stat.ee/en/stat/rahvastik_rahvastikusundmused_surmad/RV04 (accessed on 04.03.2021)

**Finland**. Statistics Finland: 12ah -- Deaths by month, 1945-2020. Available at https://pxnet2.stat.fi/PXWeb/pxweb/en/StatFin/StatFin_vrm_kuol/statfin_kuol_pxt_12ah.px/ (accessed on 06.03.2021)

**France**. INSEE, L’Institut national de la statistique et des études économiques: Demography - Number of deaths - Metropolitan France, 1946-2021. Available at https://www.insee.fr/en/statistiques/serie/000436394 (accessed on 23.02.2021)

**Hungary**. Hungarian Central Statistical Office: Main indicator of vital events (monthly data, 2017-2021). Available at http://www.ksh.hu/stadat_files/nep/en/nep0064.html (accessed on 06.03.2021)

Hungarian Central Statistical Office: Deaths in reference year (1995-2019). Available at http://statinfo.ksh.hu/Statinfo/haDetails.jsp?query=kshquery&lang=en (accessed on 06.03.2021)

**Israel**. Central Bureau of Statistics. Deaths of Israeli Residents, by Year and Month, 2000-2021 - All Ages. Data obtained by the CBS Series Generator on the topic of Population, Subtopic of Marriages, Divorces, Live-Births and Deaths. Available at https://boardsgenerator.cbs.gov.il/pages/Sdarot/wizardpage.aspx?level_1=2&level_2=2&level_3=1&level_4=1&level_5=644&l=1 (accessed on 04.03.2021)

**Italy**. ISTAT: Dataset con i decessi giornalieri in ogni singolo comune di residenza. Available at https://www.istat.it/it/archivio/240401 (accessed on 05.03.2021)

**Latvia**. Central Statistical Bureau of Latvia. IE040m. Live births and deaths by sex and by month. Available at https://data.stat.gov.lv/pxweb/en/OSP_PUB/START_POP_IM_IMSV/IDS010m (accessed on 03.03.2021)

**Lithuania**. Statistics Lithuania: Monthly and Weekly Demographic Indicators: Deaths by month. Available at https://osp.stat.gov.lt/statistiniu-rodikliu-analize#/ (accessed on 25.02.2021)

**The Netherlands**. Statistics Netherlands (CBS): Population dynamics (Deaths) by month and year. Available at https://opendata.cbs.nl/statline/#/CBS/en/dataset/83474ENG/table?ts=1623842358795 (accessed on 24.02.2021)

**New Zealand**. Stats NZ: Monthly death registrations by ethnicity-age-sex: January 2010 to December 2020. Available at https://www.stats.govt.nz/information-releases/births-and-deaths-year-ended-december-2020-including-abridged-period-life-table (accessed on 25.02.2021)

**Norway**. Statistics Norway: Deaths. Preliminary figures, by sex, year, month, age, contents and region. Available at https://www.ssb.no/en/statbank/table/12982 (accessed on 03.03.2021)

**Poland**. Statistics Poland: Statistical Bulletin No 12/2020,Tabl.7. Population and vital statistics. Available at https://stat.gov.pl/en/topics/other-studies/informations-on-socio-economic-situation/statistical-bulletin-no-122020,4,120.html (accessed on 03.03.2021)

**Portugal**. Statistics Portugal: Deaths by place of residence and sex, monthly. Available at https://www.ine.pt/xportal/xmain?xpid=INE&xpgid=ine_indicadores&indOcorrCod=0007264&contexto=bd&selTab=tab2 (accessed on 03.03.2021)

**Republic of Korea**. KOSIS: Vital statistics (deaths) by Month for Provinces. Available at https://kosis.kr/statHtml/statHtml.do?orgId=101&tblId=DT_1B8000G&language=en&conn_path=I3 (accessed on 25.02.2021)

**Scotland**. National Records of Scotland. Deaths in Scotland by month of registration and NHS Board area, 1990 – 2021. Available at https://www.nrscotland.gov.uk/statistics-and-data/statistics/statistics-by-theme/vital-events/general-publications/weekly-and-monthly-data-on-births-and-deaths/monthly-data-on-births-and-deaths-registered-in-scotland (accessed on 03.03.2021)

**Slovenia**. Republic of Slovenia Statistical Office, SIStat: Deaths by day of death, Slovenia, monthly. Available at https://pxweb.stat.si/SiStatData/pxweb/en/Data/-/05L1018S.px/ (accessed on 25.02.2021)

**Slovakia**. Statistical Office of the Slovak Republic. Deaths by Month of Death, Age, Sex and Causes of Death - SR-Area-Reg (monthly). Available at the DataCube http://datacube.statistics.sk/#!/view/en/VBD_SK_WIN2/om3801mr/v_om3801mr_00_00_00_en (accessed on 04.03.2021).

**Spain**. Instituto Nacional de Estadística: Defunciones por edad, mes y sexo. Definitivos (2010-2019) and Provisionales (2020). Available at https://www.ine.es/dyngs/INEbase/es/operacion.htm?c=Estadistica_C&cid=1254736177008&menu=resultados&idp=1254735573002 (accessed on 08.03.2021)

**Sweden**. Statistics Sweden, SCB: Deaths per month by region, Region of birth, age and sex. 2000-2020. Available at https://www.statistikdatabasen.scb.se/pxweb/en/ssd/START_BE_BE0101_BE0101I/Dod_aManadReg/ (accessed on 08.03.2021)

**Switzerland**. Office Fédéral de la Statistique: Décès par mois et mortalité depuis 1803 selon Année et Caractéristique démographique et indicateur. Available at https://www.bfs.admin.ch/asset/fr/px-x-0102020206_111 (23.02.2021)

**Taiwan.** Dept. of Household Registration Affairs, MOI. Number and Rates of Birth, Death, Marriage and Divorce. Available at https://www.ris.gov.tw/app/en/3911 (accessed on 06.03.2021)

**U.S.A**. National Center for Health Statistics, CDC-NCHS: Deaths by month tabulated from the Mortality Multiple Cause Files. https://www.cdc.gov/nchs/data_access/vitalstatsonline.htm#Mortality_Multiple (accessed on 25.02.2021)

## Appendix B Supplementary Tables

**Table 1B:** Excess mortality rates (per 100,000 person-years) for Scenarios 1-8 by country, 2020

| Country | Index | Method | Reference Period | Time Unit | Excess Mortality | Confidence Interval |  |
| --- | --- | --- | --- | --- | --- | --- | --- |
|  |  |  |  |  |  | Lower Limit | Upper Limit |
| Austria | CDR | Specific-Average | 2015-2019 | Weekly | 62.91 | 60.09 | 65.78 |
| Austria | CDR | Specific-Average with Trend | 2015-2019 | Weekly | 68.56 | 61.4 | 75.4 |
| Austria | CDR | Harmonics with Trend | 2015-2019 | Weekly | 70.36 | 63.17 | 77.08 |
| Austria | CDR | Specific-Trend | 2015-2019 | Weekly | 69.79 | 62.55 | 76.52 |
| Austria | SDR | Specific-Average | 2015-2019 | Weekly | 20.74 | 18.03 | 23.5 |
| Austria | SDR | Specific-Average with Trend | 2015-2019 | Weekly | 60.55 | 53.6 | 67.3 |
| Austria | SDR | Harmonics with Trend | 2015-2019 | Weekly | 62.15 | 55.36 | 68.91 |
| Austria | SDR | Specific-Trend | 2015-2019 | Weekly | 61.88 | 55.07 | 68.4 |
| Belgium | CDR | Specific-Average | 2015-2019 | Weekly | 142.42 | 139.83 | 145.01 |
| Belgium | CDR | Specific-Average with Trend | 2015-2019 | Weekly | 169.32 | 163.67 | 175.2 |
| Belgium | CDR | Harmonics with Trend | 2015-2019 | Weekly | 169.64 | 163.85 | 175.29 |
| Belgium | CDR | Specific-Trend | 2015-2019 | Weekly | 169.17 | 163.24 | 174.95 |
| Belgium | SDR | Specific-Average | 2015-2019 | Weekly | 90.89 | 88.46 | 93.46 |
| Belgium | SDR | Specific-Average with Trend | 2015-2019 | Weekly | 159.71 | 154.13 | 165.21 |
| Belgium | SDR | Harmonics with Trend | 2015-2019 | Weekly | 159.8 | 154.09 | 165.25 |
| Belgium | SDR | Specific-Trend | 2015-2019 | Weekly | 159.57 | 153.81 | 164.84 |
| Denmark | CDR | Specific-Average | 2015-2019 | Weekly | 5.86 | 2.23 | 9.5 |
| Denmark | CDR | Specific-Average with Trend | 2015-2019 | Weekly | -5.22 | -13.86 | 3.19 |
| Denmark | CDR | Harmonics with Trend | 2015-2019 | Weekly | -5.64 | -14.3 | 2.85 |
| Denmark | CDR | Specific-Trend | 2015-2019 | Weekly | -4.39 | -13.21 | 4.09 |
| Denmark | SDR | Specific-Average | 2015-2019 | Weekly | -42.33 | -46.01 | -38.42 |
| Denmark | SDR | Specific-Average with Trend | 2015-2019 | Weekly | -12.46 | -21.36 | -4.42 |
| Denmark | SDR | Harmonics with Trend | 2015-2019 | Weekly | -13.29 | -22.35 | -5.32 |
| Denmark | SDR | Specific-Trend | 2015-2019 | Weekly | -11.51 | -20.65 | -3.37 |
| Estonia | CDR | Specific-Average | 2015-2019 | Weekly | 23.88 | 14.84 | 32.24 |
| Estonia | CDR | Specific-Average with Trend | 2015-2019 | Weekly | 17.36 | -1.74 | 37.39 |
| Estonia | CDR | Harmonics with Trend | 2015-2019 | Weekly | 20.86 | 1.88 | 40.48 |
| Estonia | CDR | Specific-Trend | 2015-2019 | Weekly | 16.68 | -1.85 | 38.09 |
| Estonia | SDR | Specific-Average | 2015-2019 | Weekly | -37.99 | -46.38 | -29.29 |
| Estonia | SDR | Specific-Average with Trend | 2015-2019 | Weekly | 24.8 | 4.94 | 44.05 |
| Estonia | SDR | Harmonics with Trend | 2015-2019 | Weekly | 27.73 | 8.4 | 46.96 |
| Estonia | SDR | Specific-Trend | 2015-2019 | Weekly | 23.99 | 3.96 | 42.99 |
| France | CDR | Specific-Average | 2015-2019 | Weekly | 90.91 | 89.78 | 91.95 |
| France | CDR | Specific-Average with Trend | 2015-2019 | Weekly | 83.56 | 81.11 | 86.09 |
| France | CDR | Harmonics with Trend | 2015-2019 | Weekly | 85.88 | 83.45 | 88.41 |
| France | CDR | Specific-Trend | 2015-2019 | Weekly | 84.2 | 81.7 | 86.75 |
| France | SDR | Specific-Average | 2015-2019 | Weekly | 31.71 | 30.74 | 32.63 |
| France | SDR | Specific-Average with Trend | 2015-2019 | Weekly | 75 | 72.69 | 77.38 |
| France | SDR | Harmonics with Trend | 2015-2019 | Weekly | 76.76 | 74.39 | 79.14 |
| France | SDR | Specific-Trend | 2015-2019 | Weekly | 75.63 | 73.27 | 77.95 |
| Finland | CDR | Specific-Average | 2015-2019 | Weekly | 12.06 | 8.41 | 15.62 |
| Finland | CDR | Specific-Average with Trend | 2015-2019 | Weekly | -3.45 | -11.7 | 4.57 |
| Finland | CDR | Harmonics with Trend | 2015-2019 | Weekly | -0.92 | -9.27 | 7.14 |
| Finland | CDR | Specific-Trend | 2015-2019 | Weekly | -2.14 | -10.41 | 6.08 |
| Finland | SDR | Specific-Average | 2015-2019 | Weekly | -48.01 | -51.68 | -44.32 |
| Finland | SDR | Specific-Average with Trend | 2015-2019 | Weekly | -5.39 | -13.18 | 3.16 |
| Finland | SDR | Harmonics with Trend | 2015-2019 | Weekly | -3.33 | -11.01 | 5.35 |
| Finland | SDR | Specific-Trend | 2015-2019 | Weekly | -4.1 | -11.85 | 4.41 |
| Scotland | CDR | Specific-Average | 2015-2019 | Weekly | 108.93 | 105.18 | 113 |
| Scotland | CDR | Specific-Average with Trend | 2015-2019 | Weekly | 121.72 | 112 | 130.34 |
| Scotland | CDR | Harmonics with Trend | 2015-2019 | Weekly | 124.82 | 115.19 | 133.4 |
| Scotland | CDR | Specific-Trend | 2015-2019 | Weekly | 121.74 | 111.95 | 130.23 |
| Scotland | SDR | Specific-Average | 2015-2019 | Weekly | 65.52 | 61.53 | 69.24 |
| Scotland | SDR | Specific-Average with Trend | 2015-2019 | Weekly | 125.2 | 116.7 | 134.1 |
| Scotland | SDR | Harmonics with Trend | 2015-2019 | Weekly | 128.21 | 119.58 | 137.14 |
| Scotland | SDR | Specific-Trend | 2015-2019 | Weekly | 125.08 | 116.11 | 133.88 |
| England&Wales | CDR | Specific-Average | 2015-2019 | Weekly | 113.42 | 112.3 | 114.52 |
| England&Wales | CDR | Specific-Average with Trend | 2015-2019 | Weekly | 137.25 | 134.63 | 139.71 |
| England&Wales | CDR | Harmonics with Trend | 2015-2019 | Weekly | 140.14 | 137.58 | 142.64 |
| England&Wales | CDR | Specific-Trend | 2015-2019 | Weekly | 138.73 | 136.12 | 141.26 |
| England&Wales | SDR | Specific-Average | 2015-2019 | Weekly | 78.95 | 77.93 | 80.06 |
| England&Wales | SDR | Specific-Average with Trend | 2015-2019 | Weekly | 138.87 | 136.4 | 141.47 |
| England&Wales | SDR | Harmonics with Trend | 2015-2019 | Weekly | 141.75 | 139.31 | 144.32 |
| England&Wales | SDR | Specific-Trend | 2015-2019 | Weekly | 140.3 | 137.85 | 142.91 |
| Hungary | CDR | Specific-Average | 2015-2019 | Weekly | 145.2 | 141.91 | 148.14 |
| Hungary | CDR | Specific-Average with Trend | 2015-2019 | Weekly | 143.89 | 136.72 | 151.63 |
| Hungary | CDR | Harmonics with Trend | 2015-2019 | Weekly | 147.08 | 139.86 | 154.8 |
| Hungary | CDR | Specific-Trend | 2015-2019 | Weekly | 144.06 | 136.7 | 151.64 |
| Hungary | SDR | Specific-Average | 2015-2019 | Weekly | 69.33 | 66.18 | 72.63 |
| Hungary | SDR | Specific-Average with Trend | 2015-2019 | Weekly | 146.62 | 139.11 | 154.65 |
| Hungary | SDR | Harmonics with Trend | 2015-2019 | Weekly | 149.48 | 141.89 | 157.46 |
| Hungary | SDR | Specific-Trend | 2015-2019 | Weekly | 146.91 | 139.37 | 154.98 |
| Israel | CDR | Specific-Average | 2015-2019 | Weekly | 14.57 | 12.4 | 16.66 |
| Israel | CDR | Specific-Average with Trend | 2015-2019 | Weekly | 37.9 | 33.11 | 42.62 |
| Israel | CDR | Harmonics with Trend | 2015-2019 | Weekly | 39.23 | 34.2 | 43.94 |
| Israel | CDR | Specific-Trend | 2015-2019 | Weekly | 38.26 | 33.59 | 43.01 |
| Israel | SDR | Specific-Average | 2015-2019 | Weekly | -4.64 | -7.27 | -1.94 |
| Israel | SDR | Specific-Average with Trend | 2015-2019 | Weekly | 65.26 | 58.98 | 71.65 |
| Israel | SDR | Harmonics with Trend | 2015-2019 | Weekly | 67.18 | 60.83 | 73.66 |
| Israel | SDR | Specific-Trend | 2015-2019 | Weekly | 66 | 59.69 | 72.7 |
| Italy | CDR | Specific-Average | 2015-2019 | Weekly | 194.45 | 193.34 | 195.72 |
| Italy | CDR | Specific-Average with Trend | 2015-2019 | Weekly | 185.17 | 182.54 | 187.85 |
| Italy | CDR | Harmonics with Trend | 2015-2019 | Weekly | 189.64 | 187.07 | 192.27 |
| Italy | CDR | Specific-Trend | 2015-2019 | Weekly | 186.69 | 184.04 | 189.38 |
| Italy | SDR | Specific-Average | 2015-2019 | Weekly | 89.43 | 88.42 | 90.49 |
| Italy | SDR | Specific-Average with Trend | 2015-2019 | Weekly | 140.45 | 138.15 | 142.7 |
| Italy | SDR | Harmonics with Trend | 2015-2019 | Weekly | 143.66 | 141.32 | 145.94 |
| Italy | SDR | Specific-Trend | 2015-2019 | Weekly | 141.77 | 139.36 | 144 |
| Republic of Korea | CDR | Specific-Average | 2015-2019 | Weekly | 29.95 | 29.07 | 30.89 |
| Republic of Korea | CDR | Specific-Average with Trend | 2015-2019 | Weekly | 7.75 | 5.6 | 9.93 |
| Republic of Korea | CDR | Harmonics with Trend | 2015-2019 | Weekly | 8.67 | 6.51 | 10.78 |
| Republic of Korea | CDR | Specific-Trend | 2015-2019 | Weekly | 7.52 | 5.35 | 9.71 |
| Republic of Korea | SDR | Specific-Average | 2015-2019 | Weekly | -70.88 | -71.93 | -69.74 |
| Republic of Korea | SDR | Specific-Average with Trend | 2015-2019 | Weekly | 10.67 | 7.89 | 13.26 |
| Republic of Korea | SDR | Harmonics with Trend | 2015-2019 | Weekly | 11.19 | 8.37 | 13.77 |
| Republic of Korea | SDR | Specific-Trend | 2015-2019 | Weekly | 10.36 | 7.53 | 13.03 |
| Lithuania | CDR | Specific-Average | 2015-2019 | Weekly | 126.05 | 119.09 | 132.15 |
| Lithuania | CDR | Specific-Average with Trend | 2015-2019 | Weekly | 171.07 | 156.34 | 184.8 |
| Lithuania | CDR | Harmonics with Trend | 2015-2019 | Weekly | 176.34 | 161.69 | 190.32 |
| Lithuania | CDR | Specific-Trend | 2015-2019 | Weekly | 172.39 | 157.76 | 186.71 |
| Lithuania | SDR | Specific-Average | 2015-2019 | Weekly | 51.36 | 45.01 | 57.19 |
| Lithuania | SDR | Specific-Average with Trend | 2015-2019 | Weekly | 180.2 | 165.5 | 193.93 |
| Lithuania | SDR | Harmonics with Trend | 2015-2019 | Weekly | 184.8 | 170.17 | 198.37 |
| Lithuania | SDR | Specific-Trend | 2015-2019 | Weekly | 181.69 | 166.83 | 195.09 |
| Latvia | CDR | Specific-Average | 2015-2019 | Weekly | 28.16 | 19.91 | 35.53 |
| Latvia | CDR | Specific-Average with Trend | 2015-2019 | Weekly | 11.09 | -6.67 | 29.48 |
| Latvia | CDR | Harmonics with Trend | 2015-2019 | Weekly | 13.43 | -4.37 | 32.08 |
| Latvia | CDR | Specific-Trend | 2015-2019 | Weekly | 10.3 | -7.74 | 28.7 |
| Latvia | SDR | Specific-Average | 2015-2019 | Weekly | -40.61 | -48.13 | -33.25 |
| Latvia | SDR | Specific-Average with Trend | 2015-2019 | Weekly | 20.77 | 4.27 | 38.26 |
| Latvia | SDR | Harmonics with Trend | 2015-2019 | Weekly | 22.62 | 6.18 | 40.76 |
| Latvia | SDR | Specific-Trend | 2015-2019 | Weekly | 19.94 | 3.87 | 38.16 |
| Netherlands | CDR | Specific-Average | 2015-2019 | Weekly | 86.57 | 84.73 | 88.64 |
| Netherlands | CDR | Specific-Average with Trend | 2015-2019 | Weekly | 77.57 | 73.09 | 81.95 |
| Netherlands | CDR | Harmonics with Trend | 2015-2019 | Weekly | 78.5 | 73.75 | 82.93 |
| Netherlands | CDR | Specific-Trend | 2015-2019 | Weekly | 77.32 | 72.83 | 81.73 |
| Netherlands | SDR | Specific-Average | 2015-2019 | Weekly | 48.43 | 46.31 | 50.48 |
| Netherlands | SDR | Specific-Average with Trend | 2015-2019 | Weekly | 85.87 | 81.12 | 90.57 |
| Netherlands | SDR | Harmonics with Trend | 2015-2019 | Weekly | 86.64 | 81.82 | 91.23 |
| Netherlands | SDR | Specific-Trend | 2015-2019 | Weekly | 85.59 | 80.72 | 90.15 |
| Norway | CDR | Specific-Average | 2015-2019 | Weekly | -19.58 | -22.89 | -16.31 |
| Norway | CDR | Specific-Average with Trend | 2015-2019 | Weekly | -3.31 | -10.76 | 4.06 |
| Norway | CDR | Harmonics with Trend | 2015-2019 | Weekly | -1.79 | -9.05 | 5.49 |
| Norway | CDR | Specific-Trend | 2015-2019 | Weekly | -2.33 | -9.86 | 4.99 |
| Norway | SDR | Specific-Average | 2015-2019 | Weekly | -54.47 | -57.96 | -50.71 |
| Norway | SDR | Specific-Average with Trend | 2015-2019 | Weekly | -9.94 | -18.15 | -1.22 |
| Norway | SDR | Harmonics with Trend | 2015-2019 | Weekly | -8.18 | -16.75 | 0.03 |
| Norway | SDR | Specific-Trend | 2015-2019 | Weekly | -8.69 | -17.07 | -0.32 |
| New Zealand | CDR | Specific-Average | 2015-2019 | Weekly | -33.61 | -36.86 | -30.38 |
| New Zealand | CDR | Specific-Average with Trend | 2015-2019 | Weekly | -42.45 | -50.52 | -34.66 |
| New Zealand | CDR | Harmonics with Trend | 2015-2019 | Weekly | -42.77 | -50.94 | -34.86 |
| New Zealand | CDR | Specific-Trend | 2015-2019 | Weekly | -41.48 | -49.52 | -33.66 |
| New Zealand | SDR | Specific-Average | 2015-2019 | Weekly | -82.92 | -86.92 | -79.26 |
| New Zealand | SDR | Specific-Average with Trend | 2015-2019 | Weekly | -56.44 | -65.78 | -48.03 |
| New Zealand | SDR | Harmonics with Trend | 2015-2019 | Weekly | -57.04 | -66.25 | -48.77 |
| New Zealand | SDR | Specific-Trend | 2015-2019 | Weekly | -55.21 | -64.46 | -46.73 |
| Poland | CDR | Specific-Average | 2015-2019 | Weekly | 194.5 | 193.14 | 195.94 |
| Poland | CDR | Specific-Average with Trend | 2015-2019 | Weekly | 149.49 | 146.17 | 152.84 |
| Poland | CDR | Harmonics with Trend | 2015-2019 | Weekly | 151.05 | 147.64 | 154.36 |
| Poland | CDR | Specific-Trend | 2015-2019 | Weekly | 149.98 | 146.64 | 153.42 |
| Poland | SDR | Specific-Average | 2015-2019 | Weekly | 145.09 | 143.43 | 146.67 |
| Poland | SDR | Specific-Average with Trend | 2015-2019 | Weekly | 177.5 | 173.78 | 181.28 |
| Poland | SDR | Harmonics with Trend | 2015-2019 | Weekly | 178.71 | 175.02 | 182.43 |
| Poland | SDR | Specific-Trend | 2015-2019 | Weekly | 178.09 | 174.38 | 181.97 |
| Portugal | CDR | Specific-Average | 2015-2019 | Weekly | 131.41 | 128.57 | 134.29 |
| Portugal | CDR | Specific-Average with Trend | 2015-2019 | Weekly | 110.57 | 103.78 | 116.98 |
| Portugal | CDR | Harmonics with Trend | 2015-2019 | Weekly | 116.35 | 109.78 | 122.75 |
| Portugal | CDR | Specific-Trend | 2015-2019 | Weekly | 112.22 | 105.38 | 118.63 |
| Portugal | SDR | Specific-Average | 2015-2019 | Weekly | 55.07 | 52.21 | 57.69 |
| Portugal | SDR | Specific-Average with Trend | 2015-2019 | Weekly | 89.81 | 83.34 | 95.89 |
| Portugal | SDR | Harmonics with Trend | 2015-2019 | Weekly | 94.67 | 88.15 | 100.7 |
| Portugal | SDR | Specific-Trend | 2015-2019 | Weekly | 91.49 | 84.91 | 97.53 |
| Slovenia | CDR | Specific-Average | 2015-2019 | Weekly | 159.39 | 153.91 | 165.9 |
| Slovenia | CDR | Specific-Average with Trend | 2015-2019 | Weekly | 134.12 | 119.17 | 148.59 |
| Slovenia | CDR | Harmonics with Trend | 2015-2019 | Weekly | 136.56 | 121.98 | 150.82 |
| Slovenia | CDR | Specific-Trend | 2015-2019 | Weekly | 134.18 | 119.18 | 148.54 |
| Slovenia | SDR | Specific-Average | 2015-2019 | Weekly | 92.86 | 86.97 | 99.17 |
| Slovenia | SDR | Specific-Average with Trend | 2015-2019 | Weekly | 141.7 | 127.34 | 155.62 |
| Slovenia | SDR | Harmonics with Trend | 2015-2019 | Weekly | 143.71 | 129.37 | 157.86 |
| Slovenia | SDR | Specific-Trend | 2015-2019 | Weekly | 141.73 | 126.93 | 156 |
| Slovakia | CDR | Specific-Average | 2015-2019 | Weekly | 98.12 | 94.54 | 101.7 |
| Slovakia | CDR | Specific-Average with Trend | 2015-2019 | Weekly | 97.91 | 89.17 | 106.4 |
| Slovakia | CDR | Harmonics with Trend | 2015-2019 | Weekly | 100.21 | 91.72 | 108.76 |
| Slovakia | CDR | Specific-Trend | 2015-2019 | Weekly | 99.41 | 90.71 | 107.8 |
| Slovakia | SDR | Specific-Average | 2015-2019 | Weekly | 28.86 | 24.28 | 33.16 |
| Slovakia | SDR | Specific-Average with Trend | 2015-2019 | Weekly | 130.16 | 120.8 | 139.32 |
| Slovakia | SDR | Harmonics with Trend | 2015-2019 | Weekly | 132.53 | 123.39 | 141.8 |
| Slovakia | SDR | Specific-Trend | 2015-2019 | Weekly | 132.25 | 123 | 141.37 |
| Spain | CDR | Specific-Average | 2015-2019 | Weekly | 153.89 | 152.62 | 155.07 |
| Spain | CDR | Specific-Average with Trend | 2015-2019 | Weekly | 166.96 | 164.23 | 169.57 |
| Spain | CDR | Harmonics with Trend | 2015-2019 | Weekly | 170.47 | 167.77 | 173.08 |
| Spain | CDR | Specific-Trend | 2015-2019 | Weekly | 167.45 | 164.68 | 170.16 |
| Spain | SDR | Specific-Average | 2015-2019 | Weekly | 85.15 | 83.99 | 86.31 |
| Spain | SDR | Specific-Average with Trend | 2015-2019 | Weekly | 146.6 | 143.81 | 149.42 |
| Spain | SDR | Harmonics with Trend | 2015-2019 | Weekly | 149.52 | 146.72 | 152.18 |
| Spain | SDR | Specific-Trend | 2015-2019 | Weekly | 147.11 | 144.37 | 149.89 |
| Sweden | CDR | Specific-Average | 2015-2019 | Weekly | 13.34 | 10.63 | 15.96 |
| Sweden | CDR | Specific-Average with Trend | 2015-2019 | Weekly | 53.88 | 47.88 | 59.98 |
| Sweden | CDR | Harmonics with Trend | 2015-2019 | Weekly | 54.7 | 48.78 | 60.8 |
| Sweden | CDR | Specific-Trend | 2015-2019 | Weekly | 54.37 | 48.42 | 60.44 |
| Sweden | SDR | Specific-Average | 2015-2019 | Weekly | -9.62 | -12.28 | -7.12 |
| Sweden | SDR | Specific-Average with Trend | 2015-2019 | Weekly | 40.56 | 34.47 | 46.24 |
| Sweden | SDR | Harmonics with Trend | 2015-2019 | Weekly | 41.34 | 35.22 | 46.95 |
| Sweden | SDR | Specific-Trend | 2015-2019 | Weekly | 41.12 | 35.01 | 47.02 |
| Switzerland | CDR | Specific-Average | 2015-2019 | Weekly | 92.23 | 89.63 | 94.87 |
| Switzerland | CDR | Specific-Average with Trend | 2015-2019 | Weekly | 113.32 | 107.44 | 119.22 |
| Switzerland | CDR | Harmonics with Trend | 2015-2019 | Weekly | 114.24 | 108.35 | 120.16 |
| Switzerland | CDR | Specific-Trend | 2015-2019 | Weekly | 114.12 | 107.94 | 120.19 |
| Switzerland | SDR | Specific-Average | 2015-2019 | Weekly | 46 | 43.22 | 48.61 |
| Switzerland | SDR | Specific-Average with Trend | 2015-2019 | Weekly | 106.75 | 100.39 | 113.34 |
| Switzerland | SDR | Harmonics with Trend | 2015-2019 | Weekly | 107.41 | 101.1 | 114.22 |
| Switzerland | SDR | Specific-Trend | 2015-2019 | Weekly | 107.54 | 101.34 | 114.31 |
| Taiwan | CDR | Specific-Average | 2015-2019 | Weekly | -13.79 | -15.39 | -12.21 |
| Taiwan | CDR | Specific-Average with Trend | 2015-2019 | Weekly | -40.79 | -44.35 | -36.99 |
| Taiwan | CDR | Harmonics with Trend | 2015-2019 | Weekly | -40.84 | -44.48 | -37.1 |
| Taiwan | CDR | Specific-Trend | 2015-2019 | Weekly | -40.79 | -44.35 | -36.99 |
| Taiwan | SDR | Specific-Average | 2015-2019 | Weekly | -108.42 | -110.25 | -106.64 |
| Taiwan | SDR | Specific-Average with Trend | 2015-2019 | Weekly | -53.45 | -57.87 | -49.36 |
| Taiwan | SDR | Harmonics with Trend | 2015-2019 | Weekly | -54.5 | -58.79 | -50.42 |
| Taiwan | SDR | Specific-Trend | 2015-2019 | Weekly | -53.45 | -57.87 | -49.36 |
| U.S.A. | CDR | Specific-Average | 2015-2019 | Weekly | 158.1 | 157.61 | 158.53 |
| U.S.A. | CDR | Specific-Average with Trend | 2015-2019 | Weekly | 138.13 | 137.1 | 139.21 |
| U.S.A. | CDR | Harmonics with Trend | 2015-2019 | Weekly | 139.25 | 138.25 | 140.3 |
| U.S.A. | CDR | Specific-Trend | 2015-2019 | Weekly | 137.77 | 136.71 | 138.85 |
| U.S.A. | SDR | Specific-Average | 2015-2019 | Weekly | 120.25 | 119.77 | 120.73 |
| U.S.A. | SDR | Specific-Average with Trend | 2015-2019 | Weekly | 153.95 | 152.84 | 155.15 |
| U.S.A. | SDR | Harmonics with Trend | 2015-2019 | Weekly | 154.97 | 153.85 | 156.14 |
| U.S.A. | SDR | Specific-Trend | 2015-2019 | Weekly | 153.59 | 152.46 | 154.79 |

**source:** STMF-HMD (2021) and European Commission (2013).

**Table 2B:** Excess mortality rates (per 100,000 person-years) for Scenarios 2, 6, 9-12 by country, 2020

| Country | Index | Method | Reference Period | Time Unit | Excess Mortality | Confidence Interval |  |
| --- | --- | --- | --- | --- | --- | --- | --- |
|  |  |  |  |  |  | Lower Limit | Upper Limit |
| Austria | CDR | Specific-Average with Trend | 2017-2019 | Weekly | 70.27 | 60.75 | 80.27 |
| Austria | CDR | Specific-Average with Trend | 2015-2019 | Weekly | 68.56 | 61.4 | 75.4 |
| Austria | CDR | Specific-Average with Trend | 2010-2019 | Weekly | 57.82 | 53.64 | 62.36 |
| Austria | SDR | Specific-Average with Trend | 2017-2019 | Weekly | 63.99 | 53.65 | 73.6 |
| Austria | SDR | Specific-Average with Trend | 2015-2019 | Weekly | 60.55 | 53.6 | 67.3 |
| Austria | SDR | Specific-Average with Trend | 2010-2019 | Weekly | 54.98 | 50.84 | 59.3 |
| Belgium | CDR | Specific-Average with Trend | 2017-2019 | Weekly | 183.62 | 174.76 | 191.57 |
| Belgium | CDR | Specific-Average with Trend | 2015-2019 | Weekly | 169.32 | 163.67 | 175.2 |
| Belgium | CDR | Specific-Average with Trend | 2010-2019 | Weekly | 150.9 | 147.05 | 154.71 |
| Belgium | SDR | Specific-Average with Trend | 2017-2019 | Weekly | 171.42 | 163.27 | 180.06 |
| Belgium | SDR | Specific-Average with Trend | 2015-2019 | Weekly | 159.71 | 154.13 | 165.21 |
| Belgium | SDR | Specific-Average with Trend | 2010-2019 | Weekly | 142.23 | 138.41 | 146.41 |
| Denmark | CDR | Specific-Average with Trend | 2017-2019 | Weekly | -2.2 | -14.25 | 9.91 |
| Denmark | CDR | Specific-Average with Trend | 2015-2019 | Weekly | -5.22 | -13.86 | 3.19 |
| Denmark | CDR | Specific-Average with Trend | 2010-2019 | Weekly | 16.1 | 10.91 | 21.42 |
| Denmark | SDR | Specific-Average with Trend | 2017-2019 | Weekly | -7.9 | -20.19 | 3.84 |
| Denmark | SDR | Specific-Average with Trend | 2015-2019 | Weekly | -12.46 | -21.36 | -4.42 |
| Denmark | SDR | Specific-Average with Trend | 2010-2019 | Weekly | 11.91 | 6.53 | 17.39 |
| Estonia | CDR | Specific-Average with Trend | 2017-2019 | Weekly | 38.46 | 10.85 | 65.9 |
| Estonia | CDR | Specific-Average with Trend | 2015-2019 | Weekly | 17.36 | -1.74 | 37.39 |
| Estonia | CDR | Specific-Average with Trend | 2010-2019 | Weekly | 19.95 | 7.76 | 31.58 |
| Estonia | SDR | Specific-Average with Trend | 2017-2019 | Weekly | 34.78 | 6.57 | 60.45 |
| Estonia | SDR | Specific-Average with Trend | 2015-2019 | Weekly | 24.8 | 4.94 | 44.05 |
| Estonia | SDR | Specific-Average with Trend | 2010-2019 | Weekly | 39.18 | 26.33 | 52 |
| France | CDR | Specific-Average with Trend | 2017-2019 | Weekly | 99.49 | 95.99 | 102.86 |
| France | CDR | Specific-Average with Trend | 2015-2019 | Weekly | 83.56 | 81.11 | 86.09 |
| France | CDR | Specific-Average with Trend | 2010-2019 | Weekly | 72.69 | 71.22 | 74.34 |
| France | SDR | Specific-Average with Trend | 2017-2019 | Weekly | 85.24 | 81.96 | 88.54 |
| France | SDR | Specific-Average with Trend | 2015-2019 | Weekly | 75 | 72.69 | 77.38 |
| France | SDR | Specific-Average with Trend | 2010-2019 | Weekly | 66.54 | 65.14 | 68.07 |
| Finland | CDR | Specific-Average with Trend | 2017-2019 | Weekly | 7.07 | -5.79 | 19.74 |
| Finland | CDR | Specific-Average with Trend | 2015-2019 | Weekly | -3.45 | -11.7 | 4.57 |
| Finland | CDR | Specific-Average with Trend | 2010-2019 | Weekly | -1.39 | -7.19 | 4.1 |
| Finland | SDR | Specific-Average with Trend | 2017-2019 | Weekly | 5.15 | -6.8 | 17.27 |
| Finland | SDR | Specific-Average with Trend | 2015-2019 | Weekly | -5.39 | -13.18 | 3.16 |
| Finland | SDR | Specific-Average with Trend | 2010-2019 | Weekly | -3.23 | -8.67 | 2.02 |
| Scotland | CDR | Specific-Average with Trend | 2017-2019 | Weekly | 136.93 | 124.11 | 149.86 |
| Scotland | CDR | Specific-Average with Trend | 2015-2019 | Weekly | 121.72 | 112 | 130.34 |
| Scotland | CDR | Specific-Average with Trend | 2010-2019 | Weekly | 99.51 | 93.94 | 105.17 |
| Scotland | SDR | Specific-Average with Trend | 2017-2019 | Weekly | 141.4 | 128.57 | 154.73 |
| Scotland | SDR | Specific-Average with Trend | 2015-2019 | Weekly | 125.2 | 116.7 | 134.1 |
| Scotland | SDR | Specific-Average with Trend | 2010-2019 | Weekly | 99.59 | 93.81 | 105.7 |
| England&Wales | CDR | Specific-Average with Trend | 2017-2019 | Weekly | 154.45 | 150.83 | 157.98 |
| England&Wales | CDR | Specific-Average with Trend | 2015-2019 | Weekly | 137.25 | 134.63 | 139.71 |
| England&Wales | CDR | Specific-Average with Trend | 2010-2019 | Weekly | 109.84 | 108.17 | 111.47 |
| England&Wales | SDR | Specific-Average with Trend | 2017-2019 | Weekly | 156.87 | 153.21 | 160.44 |
| England&Wales | SDR | Specific-Average with Trend | 2015-2019 | Weekly | 138.87 | 136.4 | 141.47 |
| England&Wales | SDR | Specific-Average with Trend | 2010-2019 | Weekly | 109.36 | 107.64 | 111.03 |
| Hungary | CDR | Specific-Average with Trend | 2017-2019 | Weekly | 148.42 | 138.26 | 158.93 |
| Hungary | CDR | Specific-Average with Trend | 2015-2019 | Weekly | 143.89 | 136.72 | 151.63 |
| Hungary | CDR | Specific-Average with Trend | 2010-2019 | Weekly | 143.33 | 138.37 | 148.06 |
| Hungary | SDR | Specific-Average with Trend | 2017-2019 | Weekly | 150.28 | 139.78 | 161.34 |
| Hungary | SDR | Specific-Average with Trend | 2015-2019 | Weekly | 146.62 | 139.11 | 154.65 |
| Hungary | SDR | Specific-Average with Trend | 2010-2019 | Weekly | 145.75 | 141.13 | 150.56 |
| Israel | CDR | Specific-Average with Trend | 2017-2019 | Weekly | 40.5 | 33.85 | 47.81 |
| Israel | CDR | Specific-Average with Trend | 2015-2019 | Weekly | 37.9 | 33.11 | 42.62 |
| Israel | CDR | Specific-Average with Trend | 2010-2019 | Weekly | 23.92 | 20.59 | 26.97 |
| Israel | SDR | Specific-Average with Trend | 2017-2019 | Weekly | 68.88 | 60.03 | 78.14 |
| Israel | SDR | Specific-Average with Trend | 2015-2019 | Weekly | 65.26 | 58.98 | 71.65 |
| Israel | SDR | Specific-Average with Trend | 2010-2019 | Weekly | 41.68 | 37.53 | 45.71 |
| Italy | CDR | Specific-Average with Trend | 2017-2019 | Weekly | 204.41 | 200.61 | 208.31 |
| Italy | CDR | Specific-Average with Trend | 2015-2019 | Weekly | 185.17 | 182.54 | 187.85 |
| Italy | SDR | Specific-Average with Trend | 2017-2019 | Weekly | 155.42 | 151.93 | 158.77 |
| Italy | SDR | Specific-Average with Trend | 2015-2019 | Weekly | 140.45 | 138.15 | 142.7 |
| Republic of Korea | CDR | Specific-Average with Trend | 2017-2019 | Weekly | 16.53 | 13.58 | 19.55 |
| Republic of Korea | CDR | Specific-Average with Trend | 2015-2019 | Weekly | 7.75 | 5.6 | 9.93 |
| Republic of Korea | CDR | Specific-Average with Trend | 2010-2019 | Weekly | 10.23 | 8.77 | 11.54 |
| Republic of Korea | SDR | Specific-Average with Trend | 2017-2019 | Weekly | 26.79 | 23.21 | 30.44 |
| Republic of Korea | SDR | Specific-Average with Trend | 2015-2019 | Weekly | 10.67 | 7.89 | 13.26 |
| Republic of Korea | SDR | Specific-Average with Trend | 2010-2019 | Weekly | 12.73 | 11.01 | 14.42 |
| Lithuania | CDR | Specific-Average with Trend | 2017-2019 | Weekly | 189.89 | 168.96 | 209.75 |
| Lithuania | CDR | Specific-Average with Trend | 2015-2019 | Weekly | 171.07 | 156.34 | 184.8 |
| Lithuania | CDR | Specific-Average with Trend | 2010-2019 | Weekly | 116.6 | 106.79 | 126.01 |
| Lithuania | SDR | Specific-Average with Trend | 2017-2019 | Weekly | 190.57 | 169.27 | 211.51 |
| Lithuania | SDR | Specific-Average with Trend | 2015-2019 | Weekly | 180.2 | 165.5 | 193.93 |
| Lithuania | SDR | Specific-Average with Trend | 2010-2019 | Weekly | 139.73 | 129.7 | 149.91 |
| Latvia | CDR | Specific-Average with Trend | 2017-2019 | Weekly | 50.54 | 24.82 | 75.15 |
| Latvia | CDR | Specific-Average with Trend | 2015-2019 | Weekly | 11.09 | -6.67 | 29.48 |
| Latvia | CDR | Specific-Average with Trend | 2010-2019 | Weekly | 7.44 | -4.82 | 19.23 |
| Latvia | SDR | Specific-Average with Trend | 2017-2019 | Weekly | 53.23 | 27.26 | 77.78 |
| Latvia | SDR | Specific-Average with Trend | 2015-2019 | Weekly | 20.77 | 4.27 | 38.26 |
| Latvia | SDR | Specific-Average with Trend | 2010-2019 | Weekly | 32.31 | 19.93 | 43.9 |
| Netherlands | CDR | Specific-Average with Trend | 2017-2019 | Weekly | 84.33 | 77.5 | 90.67 |
| Netherlands | CDR | Specific-Average with Trend | 2015-2019 | Weekly | 77.57 | 73.09 | 81.95 |
| Netherlands | CDR | Specific-Average with Trend | 2010-2019 | Weekly | 65.07 | 61.93 | 68.11 |
| Netherlands | SDR | Specific-Average with Trend | 2017-2019 | Weekly | 92.94 | 85.54 | 100.08 |
| Netherlands | SDR | Specific-Average with Trend | 2015-2019 | Weekly | 85.87 | 81.12 | 90.57 |
| Netherlands | SDR | Specific-Average with Trend | 2010-2019 | Weekly | 78.18 | 74.86 | 81.19 |
| Norway | CDR | Specific-Average with Trend | 2017-2019 | Weekly | -1.79 | -13.31 | 9.65 |
| Norway | CDR | Specific-Average with Trend | 2015-2019 | Weekly | -3.31 | -10.76 | 4.06 |
| Norway | CDR | Specific-Average with Trend | 2010-2019 | Weekly | 10.99 | 5.83 | 16.21 |
| Norway | SDR | Specific-Average with Trend | 2017-2019 | Weekly | -5.77 | -18.18 | 5.77 |
| Norway | SDR | Specific-Average with Trend | 2015-2019 | Weekly | -9.94 | -18.15 | -1.22 |
| Norway | SDR | Specific-Average with Trend | 2010-2019 | Weekly | -2.35 | -8.28 | 3.27 |
| New Zealand | CDR | Specific-Average with Trend | 2017-2019 | Weekly | -33.54 | -45 | -23.01 |
| New Zealand | CDR | Specific-Average with Trend | 2015-2019 | Weekly | -42.45 | -50.52 | -34.66 |
| New Zealand | SDR | Specific-Average with Trend | 2017-2019 | Weekly | -44.83 | -57.02 | -33.07 |
| New Zealand | SDR | Specific-Average with Trend | 2015-2019 | Weekly | -56.44 | -65.78 | -48.03 |
| Poland | CDR | Specific-Average with Trend | 2017-2019 | Weekly | 157.15 | 152.52 | 162.49 |
| Poland | CDR | Specific-Average with Trend | 2015-2019 | Weekly | 149.49 | 146.17 | 152.84 |
| Poland | CDR | Specific-Average with Trend | 2010-2019 | Weekly | 163.39 | 161.17 | 165.73 |
| Poland | SDR | Specific-Average with Trend | 2017-2019 | Weekly | 184.55 | 179.32 | 189.91 |
| Poland | SDR | Specific-Average with Trend | 2015-2019 | Weekly | 177.5 | 173.78 | 181.28 |
| Poland | SDR | Specific-Average with Trend | 2010-2019 | Weekly | 198.51 | 196.16 | 200.98 |
| Portugal | CDR | Specific-Average with Trend | 2017-2019 | Weekly | 122.83 | 113.44 | 132.63 |
| Portugal | CDR | Specific-Average with Trend | 2015-2019 | Weekly | 110.57 | 103.78 | 116.98 |
| Portugal | CDR | Specific-Average with Trend | 2010-2019 | Weekly | 101.54 | 97.63 | 105.74 |
| Portugal | SDR | Specific-Average with Trend | 2017-2019 | Weekly | 95.31 | 85.75 | 104.47 |
| Portugal | SDR | Specific-Average with Trend | 2015-2019 | Weekly | 89.81 | 83.34 | 95.89 |
| Portugal | SDR | Specific-Average with Trend | 2010-2019 | Weekly | 95.23 | 90.77 | 99.45 |
| Slovenia | CDR | Specific-Average with Trend | 2017-2019 | Weekly | 153.91 | 133.13 | 173.24 |
| Slovenia | CDR | Specific-Average with Trend | 2015-2019 | Weekly | 134.12 | 119.17 | 148.59 |
| Slovenia | CDR | Specific-Average with Trend | 2010-2019 | Weekly | 132.74 | 123.94 | 142.16 |
| Slovenia | SDR | Specific-Average with Trend | 2017-2019 | Weekly | 156.57 | 136.84 | 175.4 |
| Slovenia | SDR | Specific-Average with Trend | 2015-2019 | Weekly | 141.7 | 127.34 | 155.62 |
| Slovenia | SDR | Specific-Average with Trend | 2010-2019 | Weekly | 145.28 | 136.23 | 154.68 |
| Slovakia | CDR | Specific-Average with Trend | 2017-2019 | Weekly | 110.33 | 97.8 | 122.77 |
| Slovakia | CDR | Specific-Average with Trend | 2015-2019 | Weekly | 97.91 | 89.17 | 106.4 |
| Slovakia | CDR | Specific-Average with Trend | 2010-2019 | Weekly | 97.78 | 91.97 | 103.33 |
| Slovakia | SDR | Specific-Average with Trend | 2017-2019 | Weekly | 154.68 | 140.27 | 168.32 |
| Slovakia | SDR | Specific-Average with Trend | 2015-2019 | Weekly | 130.16 | 120.8 | 139.32 |
| Slovakia | SDR | Specific-Average with Trend | 2010-2019 | Weekly | 125.28 | 118.01 | 132.05 |
| Spain | CDR | Specific-Average with Trend | 2017-2019 | Weekly | 191.01 | 186.89 | 194.85 |
| Spain | CDR | Specific-Average with Trend | 2015-2019 | Weekly | 166.96 | 164.23 | 169.57 |
| Spain | CDR | Specific-Average with Trend | 2010-2019 | Weekly | 133.25 | 131.52 | 134.95 |
| Spain | SDR | Specific-Average with Trend | 2017-2019 | Weekly | 163.31 | 159.34 | 167.3 |
| Spain | SDR | Specific-Average with Trend | 2015-2019 | Weekly | 146.6 | 143.81 | 149.42 |
| Spain | SDR | Specific-Average with Trend | 2010-2019 | Weekly | 127.06 | 125.27 | 128.74 |
| Sweden | CDR | Specific-Average with Trend | 2017-2019 | Weekly | 75.48 | 66.16 | 83.72 |
| Sweden | CDR | Specific-Average with Trend | 2015-2019 | Weekly | 53.88 | 47.88 | 59.98 |
| Sweden | CDR | Specific-Average with Trend | 2010-2019 | Weekly | 42.96 | 39.21 | 47.01 |
| Sweden | SDR | Specific-Average with Trend | 2017-2019 | Weekly | 64.81 | 56.05 | 73.07 |
| Sweden | SDR | Specific-Average with Trend | 2015-2019 | Weekly | 40.56 | 34.47 | 46.24 |
| Sweden | SDR | Specific-Average with Trend | 2010-2019 | Weekly | 31.35 | 27.2 | 35.3 |
| Switzerland | CDR | Specific-Average with Trend | 2017-2019 | Weekly | 115.17 | 106.63 | 124.07 |
| Switzerland | CDR | Specific-Average with Trend | 2015-2019 | Weekly | 113.32 | 107.44 | 119.22 |
| Switzerland | CDR | Specific-Average with Trend | 2010-2019 | Weekly | 100.46 | 96.23 | 104.22 |
| Switzerland | SDR | Specific-Average with Trend | 2017-2019 | Weekly | 108.17 | 99.66 | 116.59 |
| Switzerland | SDR | Specific-Average with Trend | 2015-2019 | Weekly | 106.75 | 100.39 | 113.34 |
| Switzerland | SDR | Specific-Average with Trend | 2010-2019 | Weekly | 88.84 | 84.52 | 92.69 |
| Taiwan | CDR | Specific-Average with Trend | 2017-2019 | Weekly | -35.19 | -40.4 | -29.95 |
| Taiwan | CDR | Specific-Average with Trend | 2015-2019 | Weekly | -40.79 | -44.35 | -36.99 |
| Taiwan | CDR | Specific-Average with Trend | 2010-2019 | Weekly | -50.38 | -52.67 | -47.91 |
| Taiwan | SDR | Specific-Average with Trend | 2017-2019 | Weekly | -41.46 | -48.04 | -35.97 |
| Taiwan | SDR | Specific-Average with Trend | 2015-2019 | Weekly | -53.45 | -57.87 | -49.36 |
| Taiwan | SDR | Specific-Average with Trend | 2010-2019 | Weekly | -65.41 | -68.13 | -62.7 |
| U.S.A. | CDR | Specific-Average with Trend | 2017-2019 | Weekly | 144.53 | 143.02 | 146.02 |
| U.S.A. | CDR | Specific-Average with Trend | 2015-2019 | Weekly | 138.13 | 137.1 | 139.21 |
| U.S.A. | SDR | Specific-Average with Trend | 2017-2019 | Weekly | 163.52 | 161.75 | 165.2 |
| U.S.A. | SDR | Specific-Average with Trend | 2015-2019 | Weekly | 153.95 | 152.84 | 155.15 |

**Source:** STMF-HMD (2021) and European Commission (2013).

**Table 3B:**
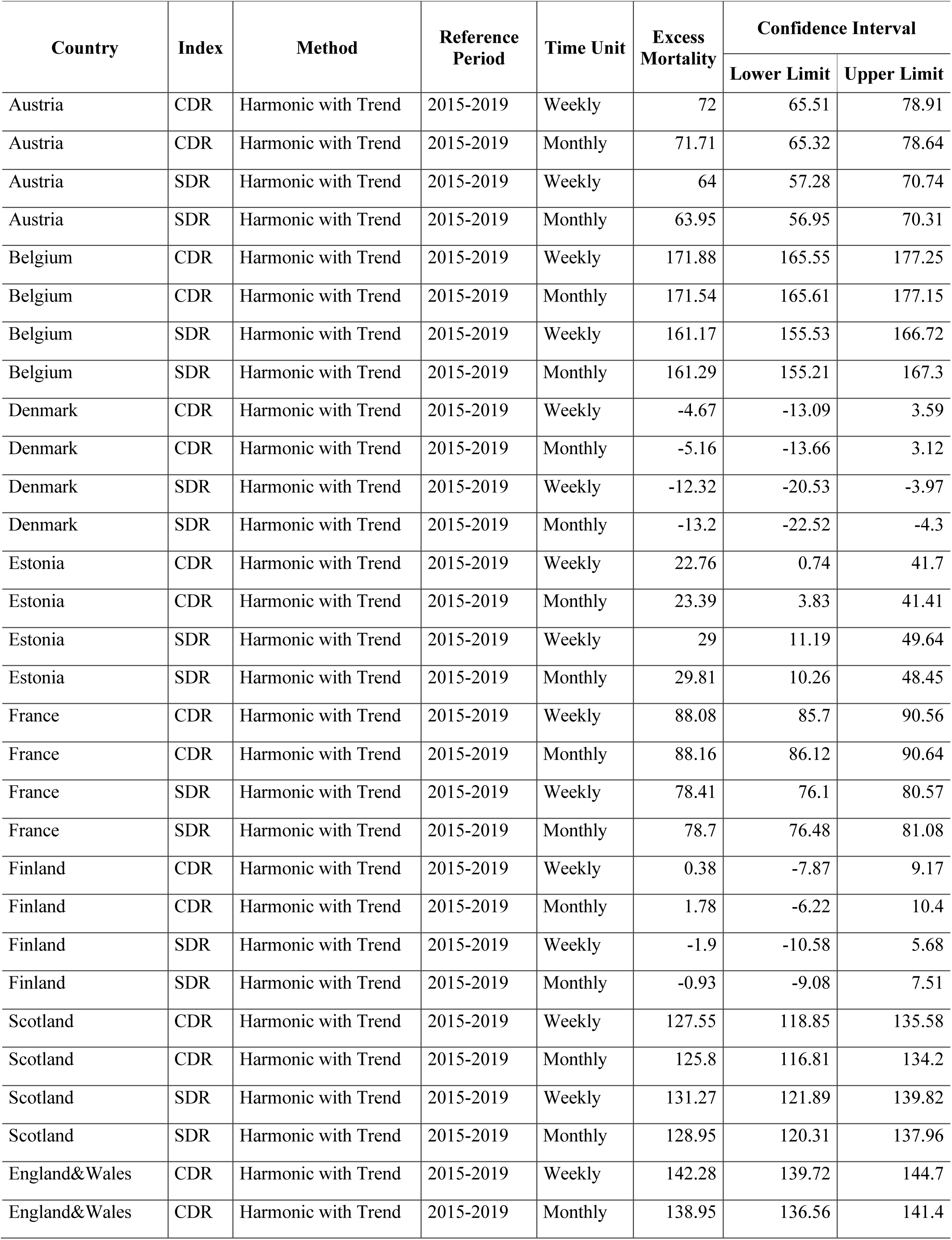
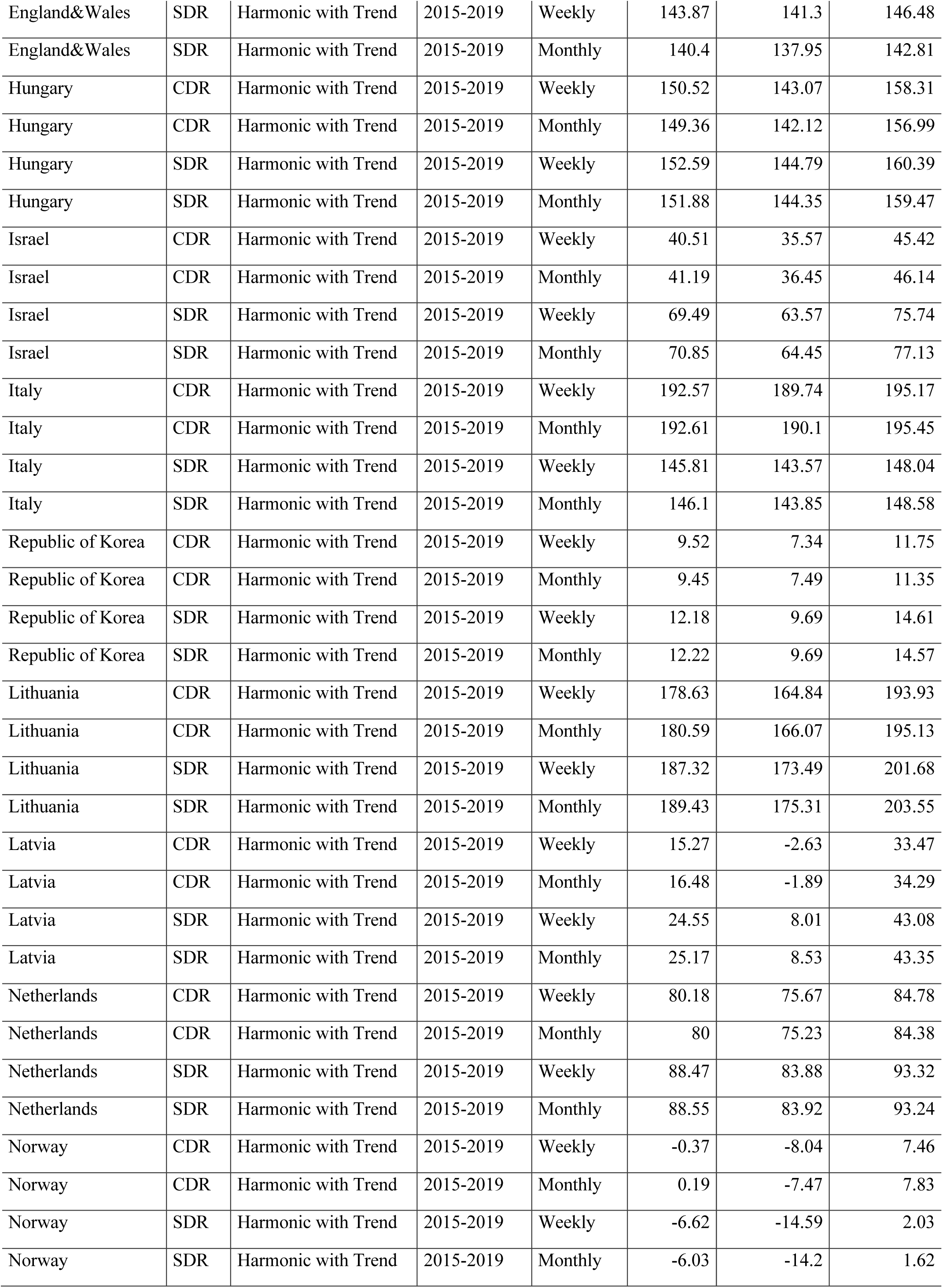
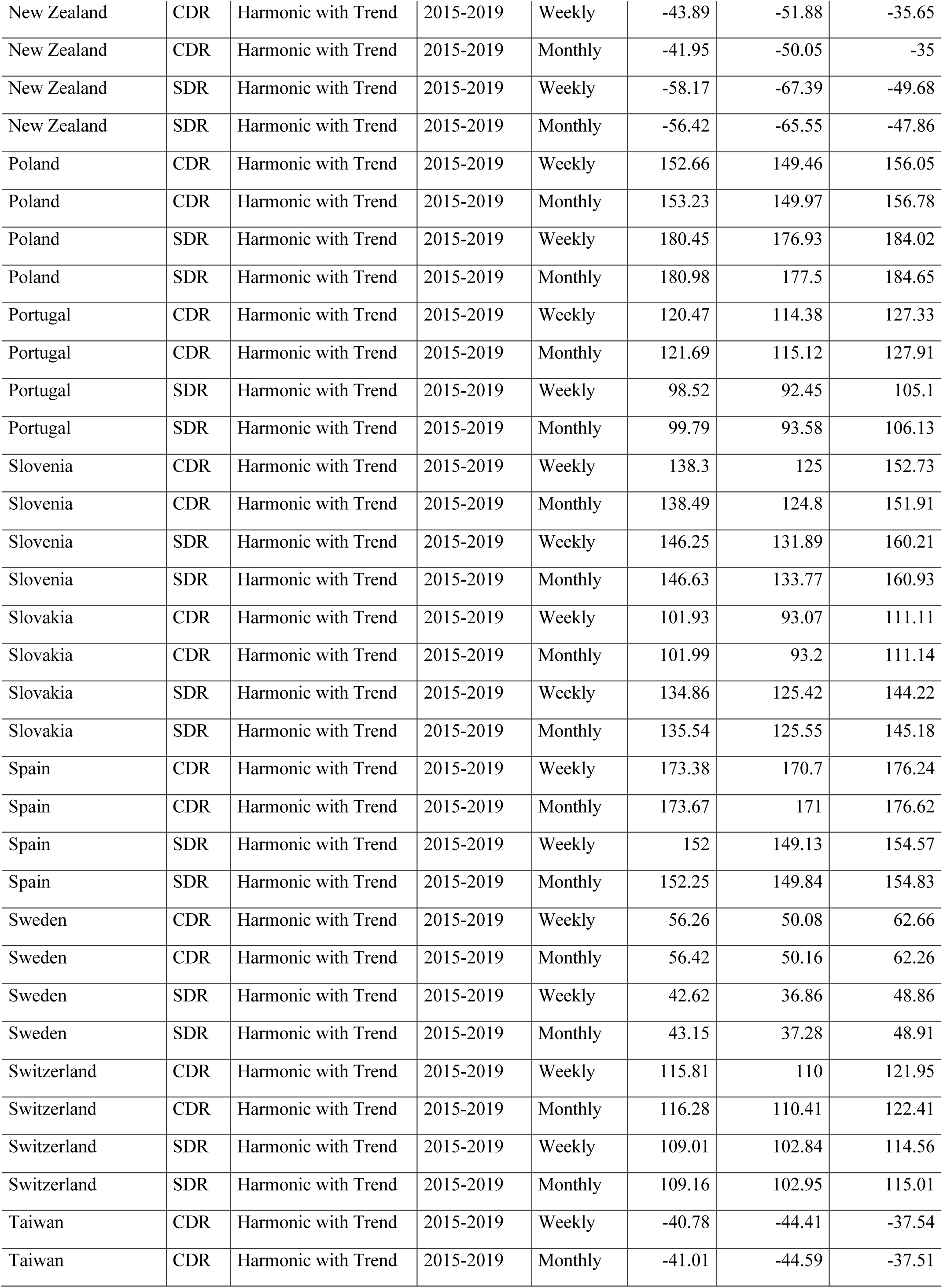
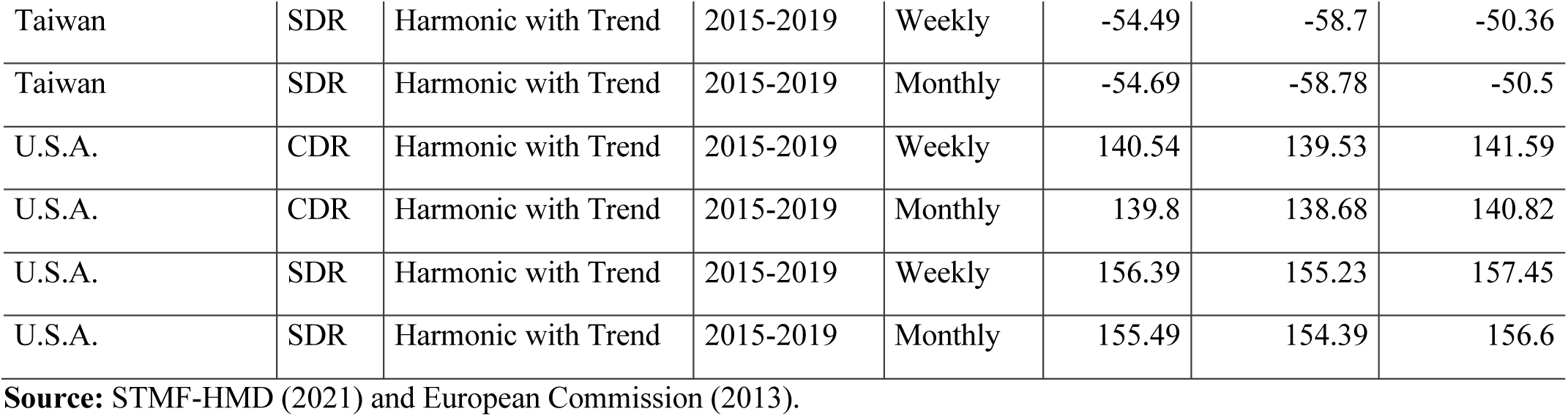
Excess mortality rates (per 100,000 person-years) for Scenarios 13-16 by country, 2020

**Table 4B:** Country ranking of excess mortality rates for each scenario, 2020

| Country | Scenario 1 | Scenario 2 | Scenario 3 | Scenario 4 | Scenario 5 | Scenario 6 | Scenario 7 | Scenario 8 | Scenario 9 | Scenario 10 | Scenario 11 | Scenario 12 |
| --- | --- | --- | --- | --- | --- | --- | --- | --- | --- | --- | --- | --- |
| Lithuania | 11 | 1 | 1 | 1 | 9 | 2 | 2 | 2 | 5 | 1 | 6 | 3 |
| Poland | 1 | 2 | 2 | 2 | 1 | 5 | 5 | 5 | 1 | 2 | 1 | 5 |
| Belgium | 4 | 3 | 3 | 3 | 7 | 3 | 4 | 3 | 4 | 3 | 2 | 4 |
| U.S.A. | 2 | 4 | 4 | 4 | 4 | 7 | 8 | 8 | - | 4 | - | 9 |
| Hungary | 8 | 5 | 6 | 6 | 6 | 6 | 6 | 6 | 2 | 10 | 3 | 8 |
| Spain | 6 | 6 | 5 | 5 | 5 | 4 | 3 | 4 | 6 | 5 | 4 | 2 |
| Slovenia | 3 | 7 | 7 | 8 | 3 | 9 | 9 | 9 | 3 | 7 | 5 | 7 |
| Italy | 5 | 8 | 8 | 7 | 2 | 1 | 1 | 1 | - | 8 | - | 1 |
| England&Wales | 7 | 9 | 9 | 9 | 10 | 8 | 7 | 7 | 8 | 6 | 7 | 6 |
| Slovakia | 15 | 10 | 10 | 10 | 12 | 13 | 13 | 13 | 7 | 9 | 11 | 13 |
| Scotland | 9 | 11 | 11 | 11 | 11 | 10 | 10 | 10 | 9 | 11 | 10 | 10 |
| Switzerland | 13 | 12 | 12 | 12 | 13 | 11 | 12 | 11 | 11 | 12 | 9 | 12 |
| Portugal | 10 | 13 | 13 | 13 | 8 | 12 | 11 | 12 | 10 | 13 | 8 | 11 |
| Netherlands | 12 | 14 | 14 | 14 | 15 | 15 | 15 | 15 | 12 | 14 | 13 | 15 |
| France | 14 | 15 | 15 | 15 | 14 | 14 | 14 | 14 | 13 | 15 | 12 | 14 |
| Israel | 17 | 16 | 16 | 16 | 20 | 18 | 18 | 18 | 15 | 16 | 16 | 19 |
| Austria | 16 | 17 | 17 | 17 | 16 | 16 | 16 | 16 | 14 | 18 | 14 | 17 |
| Sweden | 18 | 18 | 18 | 18 | 21 | 17 | 17 | 17 | 18 | 17 | 15 | 16 |
| Estonia | 19 | 19 | 19 | 19 | 19 | 19 | 19 | 19 | 16 | 20 | 17 | 20 |
| Latvia | 20 | 20 | 20 | 20 | 18 | 20 | 20 | 20 | 17 | 19 | 21 | 18 |
| Republic of Korea | 24 | 21 | 21 | 21 | 17 | 21 | 21 | 21 | 19 | 21 | 20 | 21 |
| Finland | 22 | 22 | 22 | 22 | 22 | 23 | 22 | 22 | 22 | 22 | 22 | 22 |
| Norway | 23 | 23 | 23 | 23 | 25 | 22 | 23 | 23 | 21 | 23 | 19 | 23 |
| Denmark | 21 | 24 | 24 | 24 | 23 | 24 | 24 | 24 | 20 | 24 | 18 | 24 |
| Taiwan | 26 | 25 | 25 | 25 | 24 | 25 | 25 | 25 | 23 | 25 | 23 | 26 |
| New Zealand | 25 | 26 | 26 | 26 | 26 | 26 | 26 | 26 | - | 26 | - | 25 |

Notes: Table is ordered by the Scenario 2. See Table 1 for more details about the scenarios.
Source: Author's elaboration.

## Appendix C Supplementary Figure

**Figure 1C.**
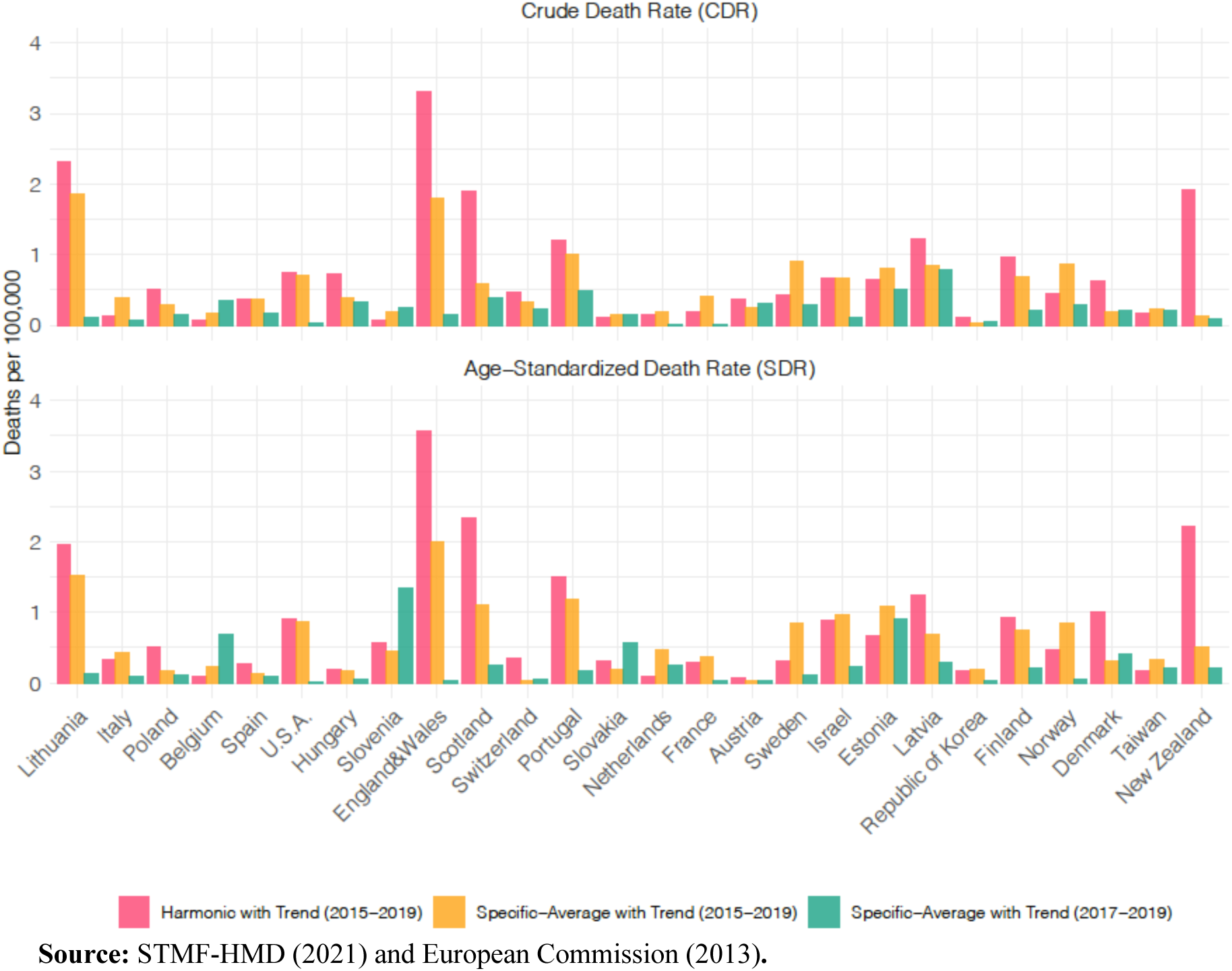
Absolute excess mortality difference between excess mortality estimates by using monthly instead of weekly data for each mortality index and country, 2020

